# Motor Module-Guided Human-Machine Interaction Promotes Improvement of Neuromuscular Coordination and Reduction in Motor Impairment in Chronic Stroke: A Pilot Study

**DOI:** 10.64898/2026.09.09.26362476

**Authors:** Manuel Portilla-Jiménez, Gang Seo, Yoon No Gregory Hong, Michael Houston, Supraja Vaidhyanathan, Yingchun Zhang, Hyung-Soon Park, Sheng Li, Jinsook Roh

## Abstract

**Background:** Stroke often leads to upper-extremity motor impairments, including abnormal neuromuscular coordination (i.e., motor modules), which reduce independent joint control and impair activities of daily living. Conventional rehabilitation mainly targets the resultant motor deficits (e.g., impaired movement) rather than the underlying neuromuscular dysfunction, potentially limiting transferability of gains beyond trained tasks. Therefore, we propose a customized, motor module-guided exercise facilitated by human-machine interaction to improve neuromuscular coordination and reduce motor impairment.

**Methods:** Five chronic stroke survivors with severe-to-mild upper extremity impairment participated in a six-week motor module-guided intervention in which they learned to selectively activate muscle groups identified in healthy individuals. In addition, three stroke survivors completed a force-guided intervention as an active control group. Standardized clinical assessments, including the Upper Extremity Fugl-Meyer Assessment and Action Research Arm Test, were performed before and after training and repeated one and three months later to assess retention. Neuromuscular coordination was quantified using non-negative matrix factorization of surface electromyography, and inter-joint coordination was evaluated during dynamic tasks using a motion capture system before and after training.

**Results:** Four of the five participants in the motor module-guided intervention improved neuromuscular coordination. These participants also showed clinically meaningful improvements in motor impairment, which were generally maintained at one and three months after training. In contrast, although the force-guided group showed improvements in task-specific performance and force control, no significant changes in neuromuscular coordination or motor impairment were observed. Notably, improvements in motor impairment correlated with enhanced stroke-induced motor module composition. Finally, findings suggest potential transferability of enhanced neuromuscular control from the trained isometric condition to untrained dynamic behaviors.

**Conclusion:** This pilot study showed that motor module-guided rehabilitation could induce improvement of stroke-affected neuromuscular coordination, even in severely impaired chronic stroke survivors, reducing motor impairment and improving motor control beyond the trained task. These findings support neuromuscular coordination-guided rehabilitation as a promising framework for post-stroke recovery.

**Trial registration:** The study was approved by the University of Houston Institutional Review Board (STUDY00001333) and registered in ClinicalTrials.gov (NCT07531264).

## INTRODUCTION

Stroke is a leading cause of long-term disability in adults, with an estimated global prevalence of 101 million cases in 2019 [1]. In the United States alone, approximately 800,000 individuals experience a stroke each year. Eighty percent of stroke survivors experience motor impairments, particularly in the upper extremity [2]. Major motor impairments, including muscle weakness, spasticity, and abnormal neuromuscular coordination, substantially reduce motor performance and limit the ability to perform activities of daily living [3]. Importantly, even after conventional rehabilitation or partial spontaneous recovery of strength and spasticity, motor impairments frequently persist [4]. Among these deficits, impaired neuromuscular coordination plays a critical role because it often leads to a loss of independent joint control and the emergence of abnormal stereotypical multi-joint movement patterns [5]. These persistent coordination deficits highlight the need for rehabilitation strategies that specifically target stroke-induced alterations in neuromuscular coordination.

Computationally identified motor modules—also referred to as muscle synergies or intermuscular coordination patterns—provide a useful proxy for characterizing alterations in neuromuscular control after stroke [6], [7], [8], [9], [10]. Here, motor modules are defined as consistent patterns of muscle coactivation across multiple muscles that are required to produce a variety of voluntary movements. This framework offers a quantitative approach to characterizing neuromuscular coordination and has strong potential to inform targeted rehabilitation interventions. Motor module analysis has been applied to investigate neuromuscular coordination of movement and force control in the upper extremity after stroke [7], [8], [11], [12], [13], [14], [15], [16]. This approach has revealed that stroke alters intermuscular coordination, as reflected by changes in the number of motor modules, their muscle composition, and their temporal activation profiles. These alterations are often associated with standardized clinical scores of motor function [17], [18], [19]. Indeed, the prevalence of abnormal neuromuscular coordination tends to increase with greater motor impairments after stroke [13], [20]. Overall, these findings indicate that motor modules provide a clinically meaningful representation of neuromuscular coordination and its impairment after stroke.

Neurophysiological evidence suggests that these abnormalities are linked to altered supraspinal control after stroke. Damage to the motor cortex and corticospinal tract can disrupt the neural orchestration of spinal motor modules, leading to characteristic changes such as module merging, fractionation, or loss [7], [21]. In addition, compensatory upregulation of descending pathways (particularly the medial reticulospinal tract, but also involved the vestibulospinal tract) and maladaptation in the spinal circuitry may contribute to abnormal stereotypical multi-joint activation patterns following stroke [22], [23], [24], [25], [26]. Studies of cortico-synergy coherence in healthy individuals further suggest that motor modules are represented at the cortical level and coordinated via alpha-band neural oscillations, supporting a cortical contribution to synergy organization [27], [28].

Beyond elucidating the neural basis of altered neuromuscular coordination, a central question for stroke rehabilitation is the extent to which these neuromuscular coordination patterns generalize across tasks, as such generalizability would suggest that targeting motor modules may extend intervention benefits beyond the specific trained task. In the lower extremity, shared motor modules have been observed across locomotor tasks such as walking and cycling in healthy individuals [29], and walking and reactive balance in stroke survivors [30]. In the upper extremity, recent studies have shown evidence of motor modules shared across distinct behaviors—such as isometric force generation and dynamic reaching—in both neurologically intact individuals [31] and stroke survivors [15]. This cross-task generalizability suggests that modifying stroke-induced motor modules through rehabilitation may yield improvements that transfer to untrained tasks including functional activities. Despite this growing body of evidence on alterations in neuromuscular coordination after stroke, the translation of motor module analysis into neurorehabilitation design remains limited.

To date, electromyogram (EMG)-guided neurorehabilitation has emerged as a promising approach to promote recovery after stroke by providing real-time information about muscle activation [32], [33]. Traditional EMG-guided rehabilitation is not motor module-guided, mostly targeting individual muscles or small muscle groups rather than multi-muscle coordination patterns [34], [35], with limited evidence for directly modulating neuromuscular coordination toward patterns identified in the healthy population. For instance, EMG-guided virtual-reality training aiming at reducing unintended wrist flexor–extensor co-activation has shown promise in promoting functional recovery and neural plasticity in individuals with severe impairment [34], [36]. Also, other EMG-guided interventions have focused on decoupling abnormal co-activation between muscle pairs (either antagonistic or agonistic pairs), resulting in reductions in motor impairment and modulation of motor module composition [37], [38], [39]. Furthermore, our previous EMG-guided study demonstrated the feasibility of directly expanding the repertoire of motor module patterns through an isometric rehabilitation paradigm designed to decouple an elbow agonist muscle pair, even in individuals with chronic and severe impairment [35].

In contrast, motor module-guided neurorehabilitation strategies could be broadly subdivided into two categories. First, some interventions use motor modules to generate stimulation patterns passively delivered to stroke survivors; the patients do not actively use motor modules in their motor performance. For instance, recent functional electrical stimulation (FES) studies used motor module patterns identified from healthy individuals to guide their FES training (also known as synergy-based FES) in the upper and lower extremities [40], [41], [42], reporting shifts toward healthier module composition alongside functional gains. Second, interventions in which motor modules are provided as real-time biofeedback require participants to actively modulate their neuromuscular coordination to achieve the training goals (e.g., the present study). To our knowledge, this last approach has not yet been investigated. Therefore, it remains unclear whether directly targeting individual motor modules can restore impaired neuromuscular coordination after stroke, reduce motor impairment, and promote transfer of training effects from practiced tasks (e.g., isometric force generation) to untrained tasks (e.g., dynamic reaching).

Thus, in this pilot study, we designed and tested a novel neuromuscular coordination-guided training protocol under isometric conditions to promote individualized recruitment of upper-extremity motor modules in chronic stroke survivors. A force-guided training was implemented as an active control to account for the potential confounding effect of repetitive isometric force generation without learning desired muscle coordination. The primary objective was to determine whether motor module-guided human–machine interaction can improve motor impairment by reducing abnormal intermuscular coordination. This work aims to advance personalized rehabilitation strategies that directly target neural recovery mechanisms after stroke.

## METHODS

### Participants

Five stroke survivors [4 males, 57 ± 11.7 years (mean ± standard deviation (SD))] were enrolled in the motor module-guided training (test group), while three stroke survivors [2 males, 53.7 ± 9.6 years] underwent the force-guided training (control group). All participants experienced a single unilateral stroke and were in the chronic stage of recovery (> 6 months since stroke onset). Upper extremity (UE) motor impairment ranged from mild to severe, as measured by the UE Fugl-Meyer Assessment (UE-FMA) (max score = 66 points). The demographics of stroke survivors are summarized in **Table 1**. The study was approved by the University of Houston Institutional Review Board (STUDY00001333), registered in ClinicalTrials.gov (NCT07531264), and conducted in accordance with the Declaration of Helsinki. Informed written consent was obtained before each assessment and training session.

**Table 1.**
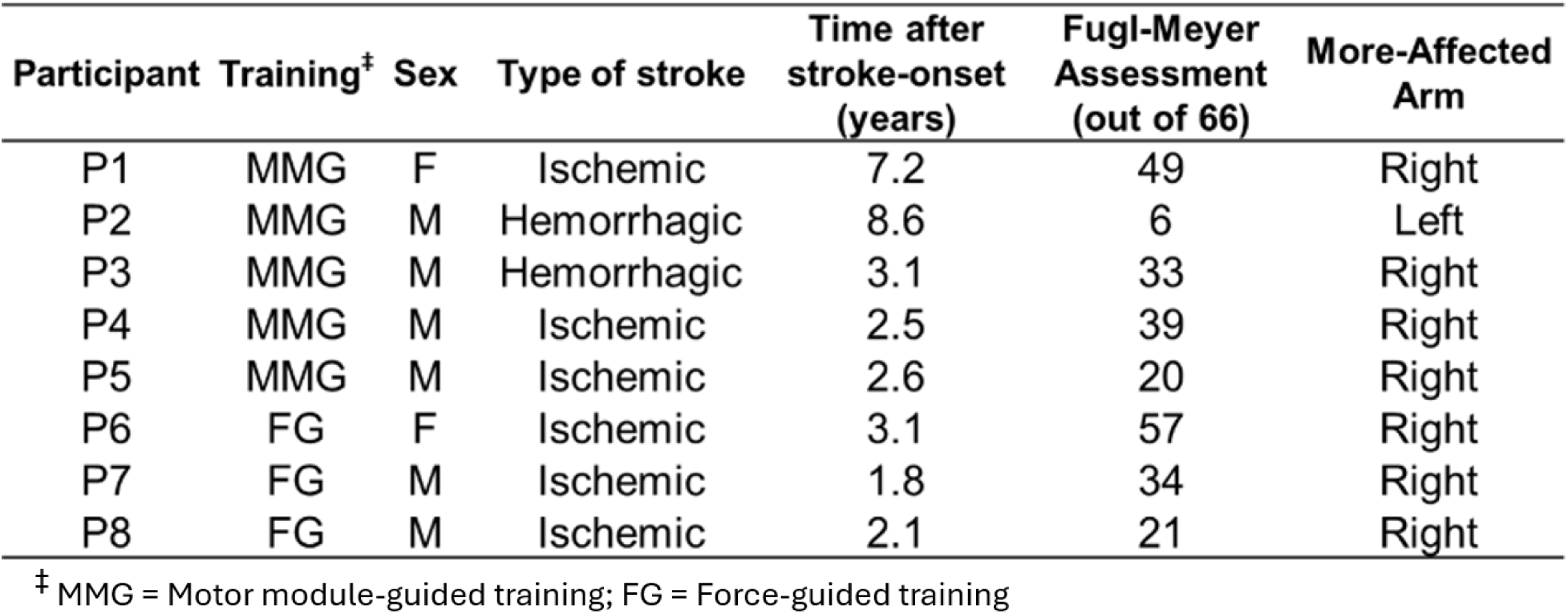
Demographic data and clinical scores.

### Equipment

Intermuscular coordination assessments and training sessions, including both motor module-guided and force-guided protocols, were conducted using a custom-designed robotic device, the KAIST Upper Limb Synergy Investigation System (KULSIS) [43]. This robotic device stabilized the arm while enabling the generation of three-dimensional forces at the hand during isometric tasks. The 3D force/torque data were measured at the hand by a six-degree-of-freedom load cell (Model: 45E15A4, JR3, Woodland, CA). Surface electromyographic (EMG) data (Trigno Wireless Biofeedback System; Delsys Inc., Boston, MA, USA) were recorded at a sampling frequency of 1 kHz.

EMG activity was recorded from eight major UE muscles: brachioradialis (BRD), biceps brachii (BB), triceps brachii (long (TrLo) and lateral (TrLa) heads), the three fibers of the deltoids (anterior (AD), middle (MD), and posterior (PD)), and the clavicular head of the pectoralis major (PEC).

For kinematic assessments, a motion capture system (Qualisys, Gothenburg, Sweden) was used to record upper limb and trunk movements. Eight reflective markers were placed on anatomical landmarks, including the acromion, incisura jugularis, the 7th cervical vertebra, 10th thoracic vertebra, lateral and medial epicondyles, and radial and ulnar styloid processes.

### Experimental Setup

For the intermuscular coordination assessments and both trainings, participants were seated with the trunk secured with a seatbelt to constrain trunk and upper-body motion and reduce compensatory movements [44]. The handle was positioned to align with the participant’s ipsilateral shoulder at a distance corresponding to 60% of his or her full arm length [13], [35], [45]. In addition, to ensure a consistent grasping posture, participants wore an auxiliary training glove.

For the training exercises, visual feedback was provided through a custom interface developed in LabVIEW, where participants interacted with a 2D display showing a circular cursor that moved based on task-related signals (i.e., multi-EMG activation or force output), depending on the training paradigm.

### Experimental Protocol

#### Designing the Motor Module-Guided Paradigm

This paradigm used motor modules as visual feedback so that post-stroke participants could train to activate desired muscle coordination. To customize the training paradigm, individualized motor module targets were defined for each participant based on their own intermuscular coordination identified from the non-paretic arm to define the targeted motor modules (e.g., **Figs. 1A and 1B**). Four commonly observed UE motor modules (see Methods section “The Procedure to Identify Motor Modules”), consistent with those previously identified in healthy individuals during a similar isometric force generation task [13], [15], [31], [45],were targeted: Elbow Flexor (EF, including BRD and BB), Elbow Extensor (EE, including TrLo and TrLa), Shoulder Flexor/Adductor (SF/Ad, including AD, MD, and PEC), and Shoulder Extensor/Abductor (SE/Ab, including MD & PD, or PD only).

**Figure 1.**
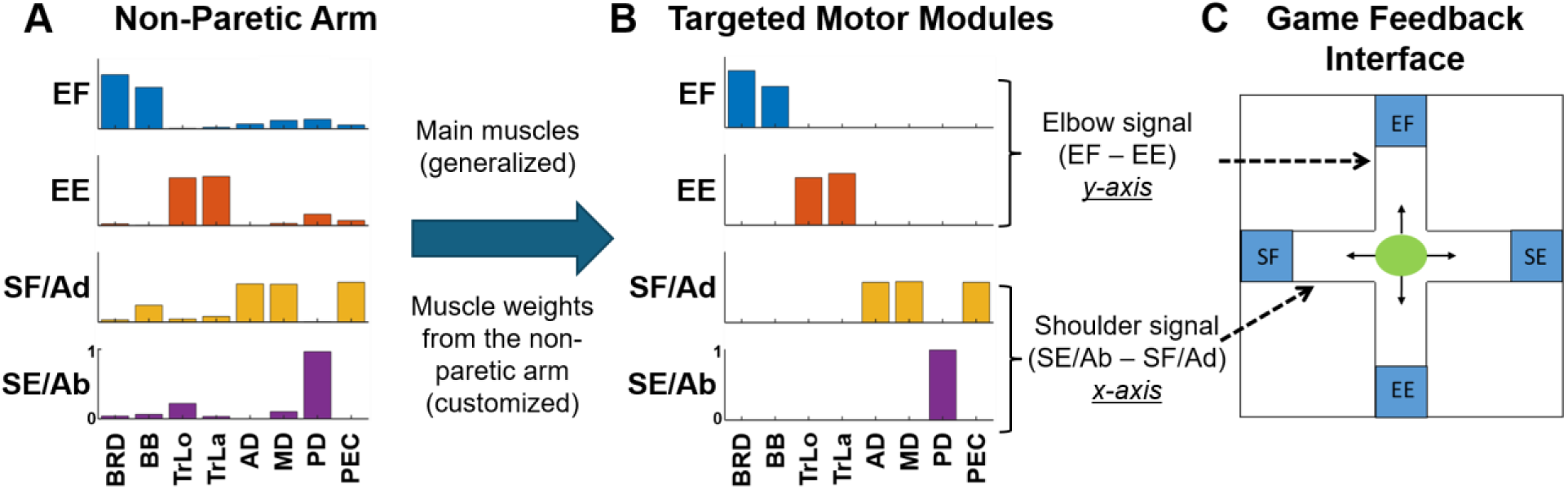
Design of individualized feedback for the motor module-guided training. Motor modules identified from the non-paretic arm of an exemplary stroke survivor **(A)**, customized targeted modules **(B)**, and game feedback interface **(C)**. Participants control the cursor (green ball) to match one of the four targets (EF, elbow flexor; EE, elbow extensor; SF/Ad, shoulder flexor/adductor; and SE/Ab, shoulder extensor/abductor) by selectively activating each motor module. The eight major UE muscles were: brachioradialis (BRD), biceps brachii (BB), triceps brachii, long (TrLo) and lateral (TrLa) heads, the three fibers of the deltoids (anterior (AD), middle (MD), and posterior (PD)), and the clavicular head of the pectoralis major (PEC).

Real-time EMG signals from the paretic arm were low-pass filtered (3rd-order Butterworth, 5 Hz as cut-off frequency). The equations that guided the cursor location during the motor module-guided training were calculated based on the predefined muscle weights from the non-paretic arm and the real-time EMG signals from the paretic arm. For instance, the two elbow motor module equations that guide the training are as follows:

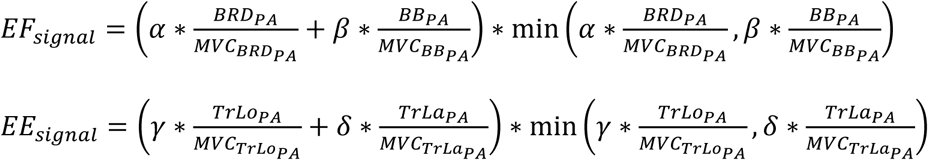

where α, β, γ, and δ are the muscle weights for BRD, BB, TrLo, and TrLa, respectively, identified from the targeted motor modules. In addition, BRD_PA_, BB_PA_, TrLo_PA_, and TrLa_PA_ are the real-time EMG signals during training from each respective muscle from the paretic arm (PA). Following the same idea, SF/Ad and SE/Ab signals were calculated to guide training. At the start of each training session, participants applied the maximal voluntary force in each of the four pre-defined motor module preferred directions (MMPD; see Methods section “The Procedure to Identify Motor Modules”) three times (12 trials, in total). These trials were used to obtain the maximum voluntary contraction (MVC) of each muscle from the paretic arm and the four maximum voluntary motor module signals (i.e., the maximum values of the four motor module equations).

The training interface was designed to provide real-time feedback based on synergistic muscle activation groups to promote selective control of individual muscle groups. During training, the four motor module equations were mapped to the displacement of the cursor in a 2D visual display in a customized LabVIEW software. EF and EE activation controlled vertical cursor motion (along the y-axis, upward and downward directions, respectively), while SF/Ad and SE/Ab activation guided horizontal cursor motion (along the x-axis, medial and lateral directions, respectively) (**Fig. 1C**).

Participants interacted with a 2D visual display where a circular cursor moved to match one of the four predefined motor module targets (EF, EE, SF/Ad, and SE/Ab). To adjust the difficulty of the training, the visual feedback display was scaled to 70% of each participant’s pre-measure maximum voluntary motor module signals. The scaling value and the margins to create the target square were selected empirically based on our previous study [35], which demonstrated that motor module characteristics could be modulated using a 70% muscle activation training paradigm, and established the margins as 15% of the maximum voluntary motor module signals. An initial feasibility assessment in the first stroke participant (P1) further supported that the task could be performed at a challenging yet achievable difficulty level. Throughout training, participants experimented with and refined their own strategies to achieve the targets, while general verbal guidance was provided when necessary to encourage activation of a single motor module at a time. For example, to reach the EE target, participants were instructed to produce an isometric force in the downward and forward direction, activating their triceps only while trying to keep the shoulder as relaxed as possible.

**Figure 2** shows four different possible scenarios for motor module-guided training. First, when a participant did not activate any muscle, the cursor remained centered on the screen, indicating a resting or baseline state (**Fig. 2A**). Second, when participants selectively activated a single synergistic muscle group, the cursor moved toward the corresponding direction on the display. For example, activating only the EF synergistic muscles (BRD and BB) caused the cursor to move upward into the target zone (top blue zone in **Fig. 2B**). Third, if two antagonistic motor modules within the same joint (e.g., EF and EE) were activated simultaneously, the signals canceled each other out, and the cursor remained near the center, failing to match the target (e.g., **Fig. 2C**). Finally, when multiple motor modules across different joints were co-activated, such as simultaneous activation of EF and SF/Ad, the cursor deviated off the intended path in a diagonal direction, missing that target (**Fig. 2D**). This interface was designed to allow participants to progressively improve their ability to isolate and control individual group of synergistic muscles, a key goal in reducing abnormal intermuscular coordination after stroke.

**Figure 2.**
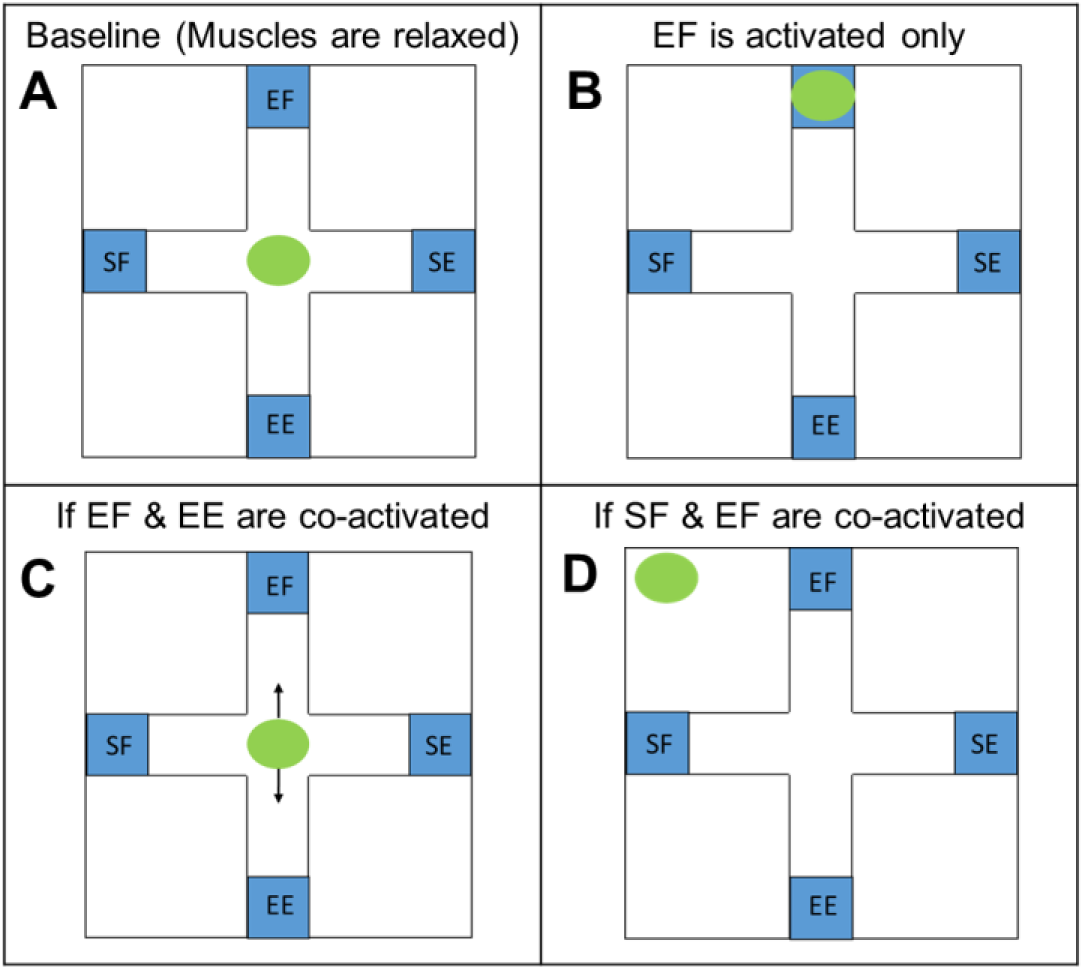
Game interface based on individualized motor module targets. Participants control the cursor, the green circle, by activating a group of muscles. **A,** A participant does not activate any muscle (baseline). **B,** If the participant activates only EF muscles, the cursor goes up to the target-match zone (blue square). **C,** If the participant activates two antagonistic groups of muscles, the target will stay close to the center; **D,** If a participant activates EF and SF motor modules, the cursor will move in a diagonal direction and be out of the target-match zone. EF, elbow flexor; EE, elbow extensor; SF, shoulder flexor/adductor; SE, shoulder extensor/abductor.

#### Designing the Force-Guided Paradigm (Active Comparator)

For the force-guided control group, the four individualized MMPDs were adopted to design the 3D target directions. Each training session began with an assessment of the four maximum voluntary forces generated in each of their respective MMPDs. Participants performed isometric strengthening exercises with force targets aligned to their MMPDs, using real-time 3D force data measured at the hand by the KULSIS system. The visual feedback display was scaled to 40% of each participant’s pre-measure maximum voluntary force in each MMPD, with a 15% logical radius to match the targets.

The 40% MVC target was selected to establish a challenging yet achievable workload that could be sustained across the entire training session. A previous study showed that even at 30% MVC, stroke survivors show greater motor unit firing rate decline on the paretic side during sustained isometric contractions compared to the non-paretic side, indicating heightened susceptibility to central fatigue [46]. In addition, our preliminary testing in healthy young adults indicated that maintaining force levels of 60–70% MVC throughout the force-guided protocol was excessively fatiguing, which is consistent with evidence that the critical torque threshold for sustainable intermittent isometric contractions lies at approximately 35–55% MVC in healthy individuals, above which neuromuscular fatigue develops rapidly [47].

#### Training Protocol for Both Exercises

To match a target in the motor module-guided training, participants received real-time EMG-based feedback (**Fig. 3A**), and a synergistic group of muscles should be activated while relaxing the other muscles to successfully match a target. Meanwhile, the force-guided group focused solely on endpoint force control (**Fig. 3B**), and the force in a specific 3D direction should be generated to successfully match a target. **Figures 3C** and **3D** show examples of successful target matches during the motor module-guided and force-guided exercises, respectively. During training, the four targets were presented in a pseudorandom order. Each trial included a 3-second inter-trial interval, a 2-second baseline, and up to 9 seconds to match a target successfully. To successfully match a target, the participant should move the cursor to the target match area and hold it there for 1 second. If a target was not matched, an additional attempt in the same direction appeared, and a maximum of three attempts were allowed per target repetition. Both training exercises consisted of one hour of training per session, three days per week for six weeks (18 training sessions in total). Each training session consisted of three training blocks (approximately 15 min each; about 60 attempts per block) separated by a break (at least 5 minutes).

**Figure 3.**
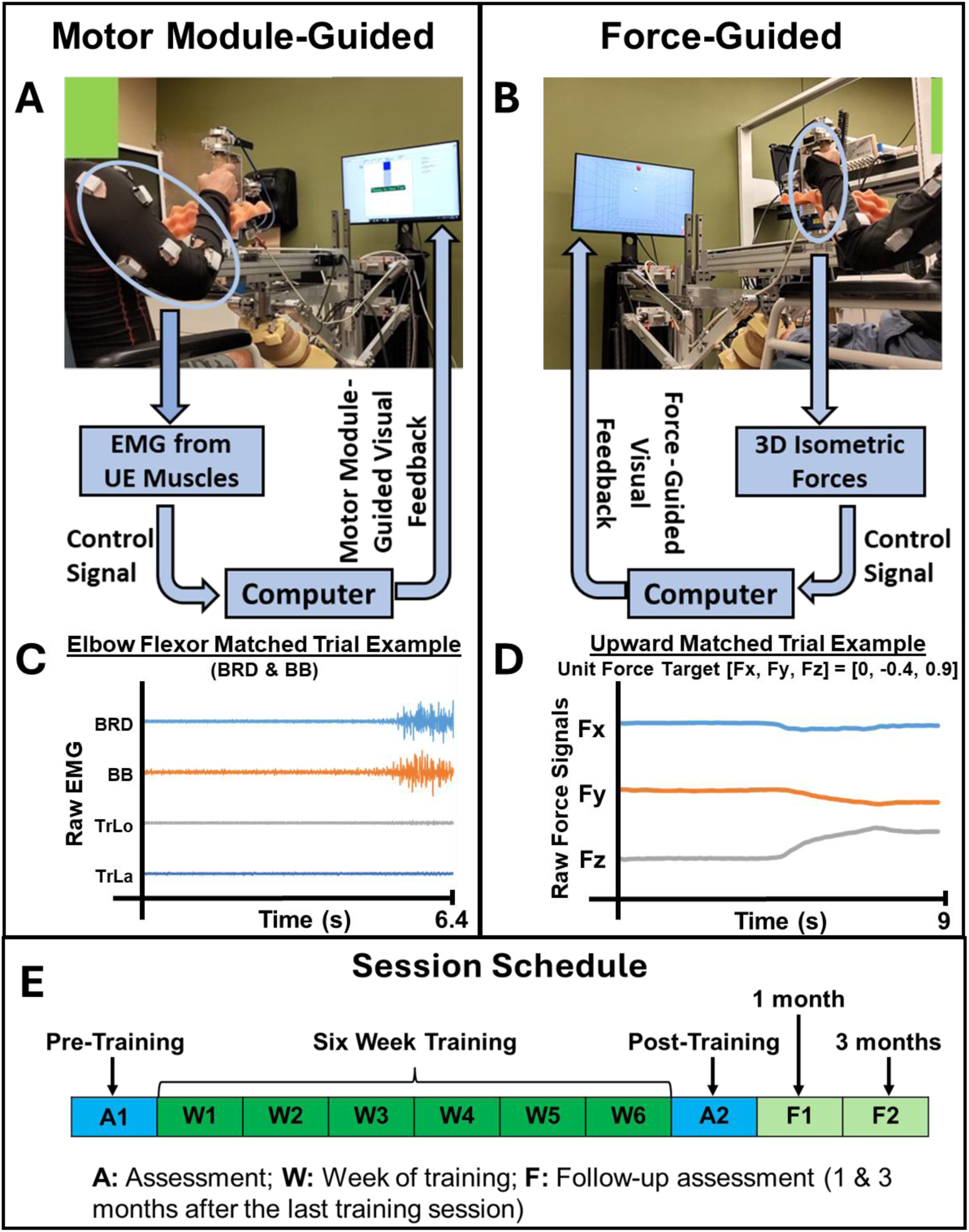
Setups of the two exercises, which adopt human-machine interaction in isometric force generation. A, Motor module-guided exercise provides multiple EMG activations as visual feedback, and a selective, desired muscle coordination pattern is required for target matches. B, Force-guided exercise provides 3D forces as visual feedback; as long as the target force is achieved, any muscle activation enables target matches. Two examples of matched trials: elbow flexor trial during motor module-guided exercise (C), and upward trial during force-guided exercise (D). Session schedule for both exercises (E). Brachioradialis (BRD), biceps brachii (BB), triceps brachii, long (TrLo) and lateral (TrLa) heads.

One participant (P2) with severe motor impairment (pre-training UE-FMA = 6 out of 66) was unable to generate and sustain 70% of maximum voluntary motor module signals for the required one-second hold during the first 3 weeks of training. Therefore, the parameters were adapted to reduce the difficulty, so successful target matches were defined as 40% of his pre-measure maximum voluntary motor module signals, and maintaining it for 0.5 s. In addition, he performed 9 additional training sessions (27 training sessions, in total) because of the modification of the game difficulty.

The session schedule consisted of four assessment sessions and 18 training sessions (**Fig. 3E**). Standardized clinical assessments, including the UE Fugl Meyer Assessment (UE-FMA) for motor impairment and the Action Research Arm Test (ARAT) for motor function, were performed by a certified physical therapist before and after training and at one and three months after the last training session. UE-FMA served as the primary outcome measure. In addition, at baseline and post-training assessments, intermuscular coordination was identified using motor module analysis. Finally, kinematic assessment and intermuscular coordination analysis underlying untrained, dynamic conditions were used to investigate potential transferability from the trained task (i.e., isometric condition) to an untrained task (i.e., dynamic condition).

#### Intermuscular Coordination Assessment (Underlying Isometric Task)

To quantify intermuscular coordination, motor module identification was conducted using surface EMG signals collected during isometric force-matching tasks in 54 directions, evenly distributed in 3D space [12], [13], [35], [45]. This large set of directions was used to identify a robust set of motor modules that explain the potentially maximum variance of EMG activation in the eight muscles generated in the 3D UE workspace. Each participant’s paretic arm was tested both pre- and post-training to investigate training-related changes. Also, the non-paretic arm was assessed only at baseline to obtain the targeted motor module characteristics. At the beginning of the assessment, each participant’s maximum lateral force (MLF) was measured with the hand positioned at the initial arm posture (i.e., holding the handle at 60% of the participant’s full arm length from the ipsilateral shoulder and the hand at the level of the glenohumeral joint). To reduce compensatory movements during this assessment, the following strategies were applied: (1) participants wore a seatbelt to constrain trunk and upper-body motion; (2) they received verbal instructions to complete the task without moving the arm and trunk; (3) compensatory movements were visually monitored during the intermuscular coordination assessments, and real-time verbal feedback was provided as needed by the operator; and (4) all participants performed practice trials, which helped them reduce arm and trunk movement as much as possible. At the beginning of the assessment, the KULSIS’s handle supported the participant’s relaxed arm against gravity. Then, the load cell attached to the handle was zeroed to remove any force signals attributable to the arm’s weight, allowing for precise measurement of three-dimensional forces. The goal of the assessment was for participants to apply and maintain a force equal to 40% of their MLF output for one second in each of the 54-targets randomly displayed [12], [13].

#### Post-Training Intermuscular Coordination Assessment (Underlying Dynamic Task)

Post-training intermuscular coordination underlying a dynamic task was assessed using surface EMG from the same eight upper-extremity muscles recorded during a point-to-point reaching task performed in horizontal and frontal planes (12 targets per plane, evenly distributed; 24 reaching targets in total). The number of targets was selected to ensure robust extraction of motor modules, as previous studies have shown that at least six reaching directions are required to obtain valid muscle synergies [48]. For both exercises, post-training motor modules from the paretic arm were compared with motor modules identified from healthy individuals in our previous study [15], whose data set served as the reference for this analysis. Detailed methods for this assessment are described in our previous study [15].

#### Kinematic Assessment

At pre- and post-training for the mildly and moderately impaired participants, inter-joint coordination during dynamic tasks was assessed using the data obtained from point-to-point reaching and drinking tasks [49]. For the reaching task, four diagonal directions on the horizontal plane were selected to capture distinct joint coordination patterns. The starting hand position was located along the trunk midline at a distance equal to 70% of the arm length. Targets were placed 15cm from the starting position. Participants completed five repetitions per direction at a self-selected speed. Since participants with severe impairment could not perform this unsupported reaching and drinking tasks, they were assessed using full active range of motion (ARoM) tasks for shoulder flexion/extension, internal/external rotation, abduction/adduction, elbow flexion/extension.

### Data Analysis

#### The Procedure to Identify Motor Modules

The raw EMG signals recorded from the eight UE muscles during the intermuscular coordination assessment underlying the isometric condition were processed using customized MATLAB software. Initially, electrocardiogram (ECG) artifacts were detected through visual inspection and subsequently removed using a wavelet-based filtering method. Next, the data were demeaned to remove the DC offset, and a full-wave rectification was applied. Then, baseline subtraction reduced noise. Lastly, to extract the EMG envelope, a low-pass filter was applied using a 4th-order Butterworth filter with a cutoff frequency of 10 Hz. The EMG data during the one-second holding period from all trials were concatenated for each assessment. Each muscle’s EMG data were normalized to unit variance to prevent dominance from muscles with higher signal variability [12], [31], [45].

Each participant’s pre-processed EMG data were analyzed for each arm separately using non-negative matrix factorization (NNMF) to identify motor modules. To satisfy the NNMF’s non-negativity constraint, all negative preprocessed EMG values were set to zero. The eight-arm muscles’ EMGs were represented as a linear combination of a set of time-invariant motor modules (W) and their respective time-varying activation profiles (C) as follows:

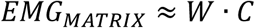

where W is an M (number of muscles = 8) by N (number of motor modules) matrix, and C is an N by D (number of data sample points) matrix. To determine the minimum number of motor modules required to accurately reconstruct the spatial characteristics of the EMG signals, the variance accounted for (VAF) was calculated across the entire dataset, referred to as the global VAF (gVAF). VAF was defined using the following formula.

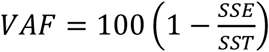

where SSE is the sum of the squared residuals, and STT is the summation of the squared EMG data [13]. The optimal number of motor modules was selected based on three criteria: 1) global variance accounted for (gVAF) exceeding 90%; 2) the addition of a new motor module contributing at least 5% additional gVAF; and 3) the VAF for each muscle greater than 60% [31], [50].

Motor modules were identified separately from the paretic arm during pre- and post-training isometric assessments. Additionally, the MMPD in 3D force space was estimated by the linear summation of the motor module activation profile as a function of the target force vector identified from the paretic arm.

#### Calculation of Motor Module Similarity

Similarity indices between motor modules from the paretic and the targeted motor modules were quantified using the scalar (dot) product between the best-matched pairs of motor module vectors across all possible combinations within each participant [7]. To estimate the similarity threshold, 1000 random sets of motor module patterns were generated by randomly selecting muscle weights from the identified motor modules across all participants and both exercises. Each vector was normalized by dividing by its magnitude to obtain unit vectors. Dot products were then calculated for all possible pairwise combinations of these motor module vectors, sorted in ascending order, and the threshold was defined as the 95th percentile of the resulting distribution [13], [15], [31], resulting in a value of 0.822. Motor modules underlying isometric condition were considered statistically similar (*p* < 0.05) if their scalar product exceeded this threshold.

Pre-training motor modules from the paretic arm were individually compared with all targeted motor modules identified from the non-paretic arm. A module was considered preserved if its maximum scalar product with any motor module from the non-paretic arm exceeded the threshold (0.822), and altered otherwise, suggesting abnormal stroke-induced motor module composition. Next, motor module similarity was calculated between each post-training altered motor module and its respective targeted motor module. Pre-training motor modules classified as preserved were excluded from this analysis, as the focus was on training-related changes in abnormal motor modules only. The difference between the post-training and pre-training similarity indices was then computed and averaged within each participant to quantify changes in altered intermuscular coordination resulting from training.

Finally, Spearman’s rank correlation was used to assess monotonic relationships between changes in altered intermuscular coordination and changes in UE-FMA after six weeks of training. Significance for the Spearman correlation analysis was set at p < 0.05.

#### Intermuscular Coordination Analysis Underlying Dynamic Task

Following our previous study procedure [15], EMG signals were preprocessed (artifact removal, full-wave rectification, and filtering) and normalized to unit variance to prevent any bias toward muscles with higher variance in the motor module identification procedure [51]. NNMF was applied to identify post-training motor modules from the paretic arm. Post-stroke survivors typically required four motor modules to adequately reconstruct upper extremity muscle activity during point-to-point reaching, whereas healthy controls required five [15]. To enable direct pairwise comparison of synergy structure between groups, the number of motor modules extracted from the healthy group was set to four, matching the stroke group. Reference (norm) motor modules were obtained by averaging and normalizing the motor modules across the eight healthy participants from our previous study [15]. Each motor module was labeled according to the primary mechanical action of its dominant muscles: elbow flexor (EF), elbow extensor (EE), shoulder flexor/adductor (SF/Ad), and shoulder extensor/abductor (SE/Ab).

To quantify the similarity between each stroke survivor’s motor modules and the healthy reference set, the scalar product was computed between each post-stroke motor module and all four reference motor modules. The similarity threshold for the point-to-point reaching task was determined using the same procedure described for the isometric analysis (see Methods section “Calculation of Motor Module Similarity”), but using motor module vectors identified from the dynamic task, yielding a threshold underlying the dynamic condition of approximately 0.814 (p < 0.05). A motor module from stroke survivors was considered statistically similar to its corresponding healthy reference module if the highest scalar product across all pairwise comparisons exceeded this threshold.

#### Kinematic Analyses

The locations of eight reflective markers were used to estimate the coordinate system of the trunk, clavicle, humerus, and forearm segments. Marker trajectory data were filtered using a fourth-order, low-pass Butterworth filter (3 Hz cutoff). These filtered trajectories were then used to estimate the coordinate system of each body segment, adopting the definitions from previous studies [52]. Joint kinematics was calculated using the rotation sequence recommended by the International Society of Biomechanics [53].

Changes in inter-joint coordination were assessed in two ways: 1) kinematic synergy similarity score [54] and 2) pairwise joint angle-to-angle correlation (e.g., between shoulder abduction and elbow flexion). Kinematic synergies were identified using NNMF [55]. Since NNMF requires positive constraints, the degrees of freedom at each joint were separated into two distinct motions (e.g., elbow flexion/extension was divided into separate elbow flexion and extension components) [56]. Kinematic synergy similarity between stroke survivors and healthy participants was calculated using their scalar product. Pairwise joint angle-to-angle correlations were calculated using Pearson’s correlation coefficient between joint angles during the point-to-point reaching task.

### Additional Statistical Analyses

The following variables did not satisfy the normality assumption based on the Shapiro–Wilk test: 1) the number of targets matched per set, comparing the first week and the last week; and 2) the average target-matching time per participant. As a result, non-parametric statistical methods were used, with the Wilcoxon signed-rank test applied for paired comparisons. All statistical significance was defined as p < 0.05.

## RESULTS

### Improvements in Task Performance during Training

Over the course of the six-week training period, both the motor module-guided and force-guided groups showed improvements in training task performance, as reflected by an increased number of successfully matched targets per set during the last week of training compared with the first week (**Fig. 4A**). In the motor module-guided group, all five stroke participants showed a statistically significant increase in the number of matched targets using a one-tailed Wilcoxon signed-rank test on paired block-wise comparisons (*, p < 0.05; **, p < 0.01). By the last week of training, participants were generally able to match targets across all four directions. One exception was P2, the participant with severe motor impairment, who was unable to successfully match the Shoulder Flexion/Adduction (SF/Ad) target during the six-week training period, although he later developed a strategy to match this target by the 27th session. Participants in the force-guided group similarly showed also increases in the number of matched targets per set from the first to the last week of training. Time taken to match targets also changed across training (**Fig. 4B**). These time results should be interpreted together with the success data, particularly for cases such as P2, where the first-week value reflects no successful matches rather than faster target matching, and the later nonzero value reflects the emergence of successful target matches.

**Figure 4.**
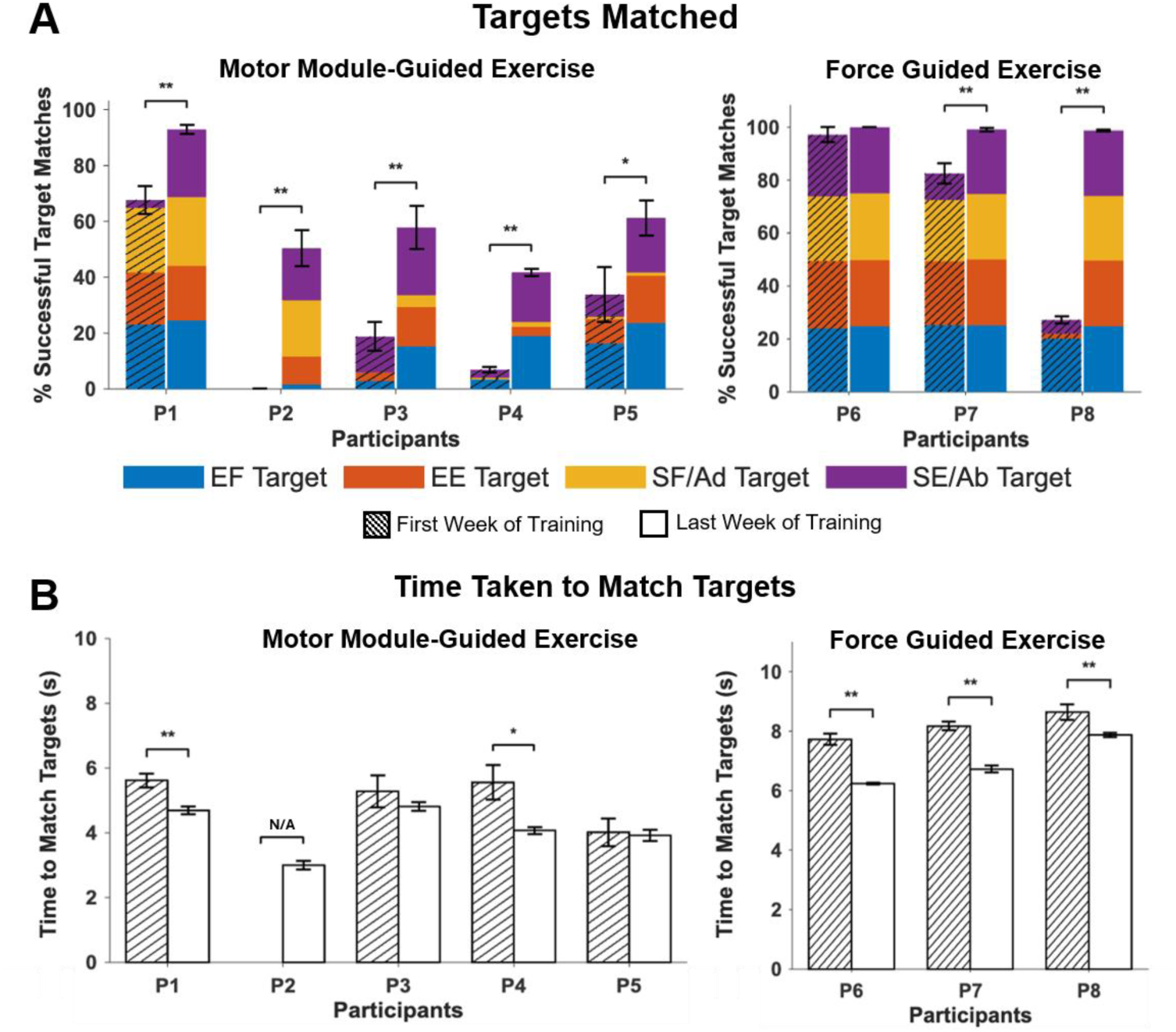
Comparison of task performance between the first and last week of training in the motor module-guided and force-guided groups. **A**, The percentage (mean ± SD) of matched targets per set for the motor module-guided group (P1–P5) and the force-guided group (P6–P8). Stacked bars indicate performance across four target directions color-coded: Elbow Flexion (EF) in blue, Elbow Extension (EE) in red, Shoulder Flexion/Adduction (SF/Ad) in orange, and Shoulder Extension/Abduction (SE/Ab) in purple. **B**, The time (mean ± standard error) taken to match targets per participant. Hatched bars represent the first week of training, while white bars represent the last week of training. *, p < 0.05; **, p < 0.01). N/A, not applicable.

### Effects of Two Exercises on Motor Impairment and Neuromuscular Coordination

#### Clinical Assessments

Four out of five participants with mild to severe motor impairment exhibited a reduction in motor impairment following motor module-guided training, as assessed by the UE-FMA (**Fig. 5A**). These four stroke survivors improved by an average of 9.25 points, exceeding the Minimal Clinically Important Difference (MCID ≥ 4) for the UE-FMA [57]. The other stroke survivor (P5) did not improve motor impairment after the motor module-guided training, and since the study’s primary outcome was UE-FMA, P5 was classified as a non-responder. Notably, the improvements in the four responders were typically maintained after 1 and 3 months of the last training session. For the participant with severe motor impairment (P2) who performed three additional weeks of training (9 weeks of training in total), the UE-FMA score was 12 at the end of 6^th^ Week.

**Figure 5.**
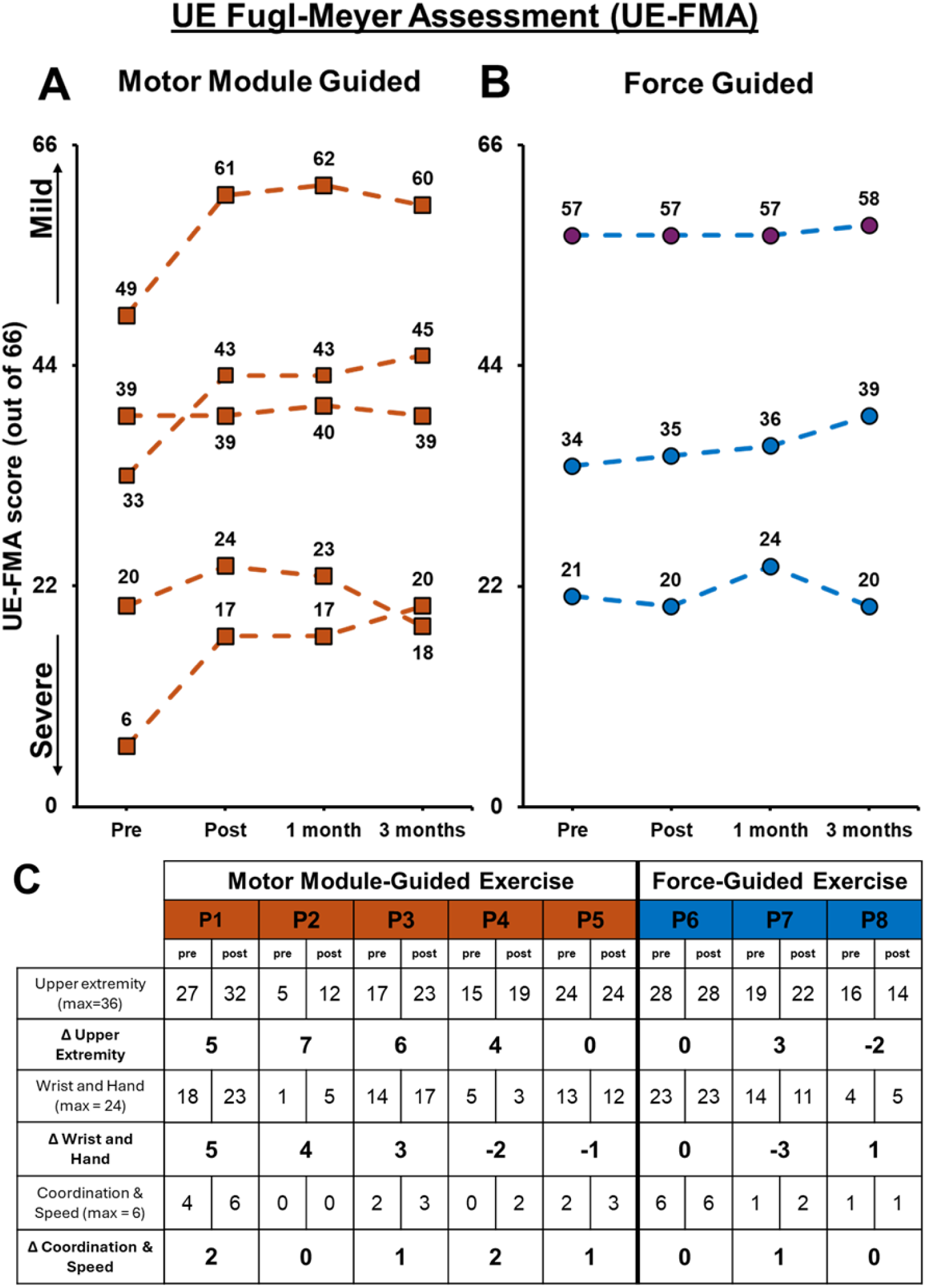
Upper extremity Fugl-Meyer assessment (UE-FMA) pre- and post-training, and one month and three months after the last training session for both motor module-guided **(A)** and force-guided **(B)** exercises. **C,** Table to detail the UE-FMA values in both exercises. Pre, pre-training; post, post-training.

In contrast, the force-guided group did not show an increase in the FMA score above MCID after the intervention. P6 remained at 57, P7 increased minimally from 34 to 35, and P8 declined slightly from 21 to 20 (**Fig. 5B**). **Figure 5C** showed detailed changes in UE-FMA subsections in both exercises. In the motor module-guided group, four out of five cases improved the scores of the FMA subsection for the upper extremity, the wrist and hand, as well as the coordination and speed, except one non-responder. The amount of the change was not observed in the force-guided group after the six weeks of training.

The Action Research Arm Test (ARAT) further assessed upper extremity functional performance. In the motor module-guided group, two of the four UE-FMA responders showed improvements in ARAT scores, with P3 improving by 10 points and P4 by 4 points, whereas P1 and P2 showed no change (**Supplementary Fig. 1**). In the force-guided group, P6 and P8 each improved by 3 points, while P7 showed a slight decrease of 2 points.

#### Neuromuscular Coordination

The motor module-guided feedback induced changes in the characteristics of motor modules in stroke, but force-guided feedback did not. The number of motor modules in the paretic arm did not change after both interventions (typically four across all participants). However, improvements in altered motor module composition were observed exclusively in the motor module-guided group (**Supplementary Fig. 2**). **Figure 6** shows two illustrative participants’ pre- and post-training motor module patterns from each group. Before training, the participant with mild motor impairment (P1; pre-training UE-FMA = 49 out of 66) in the motor module-guided training exhibited abnormal co-activation of elbow flexors and shoulder adductor/flexor muscles within the EF motor module (blue pre-training module in **Fig. 6A**). Also, abnormal coupling of all three deltoid fibers with the pectoralis major was observed within the SF/Ad motor module (orange pre-training module in **Fig. 6A**). After six weeks of motor module-guided exercise, these abnormal patterns were improved and resembled the targeted modules identified from the non-paretic arm (**Fig. 6B**), showing a clear shift toward the intended intermuscular coordination targets. Overall, four out of the five participants (P1, P2, P3, and P4) showed changes in intermuscular coordination following motor module-guided training, whereas one non-responder (P5) showed neither changes in motor module composition (**Supplementary Fig. 3)** nor improvement in the UE-FMA score.

**Figure 6.**
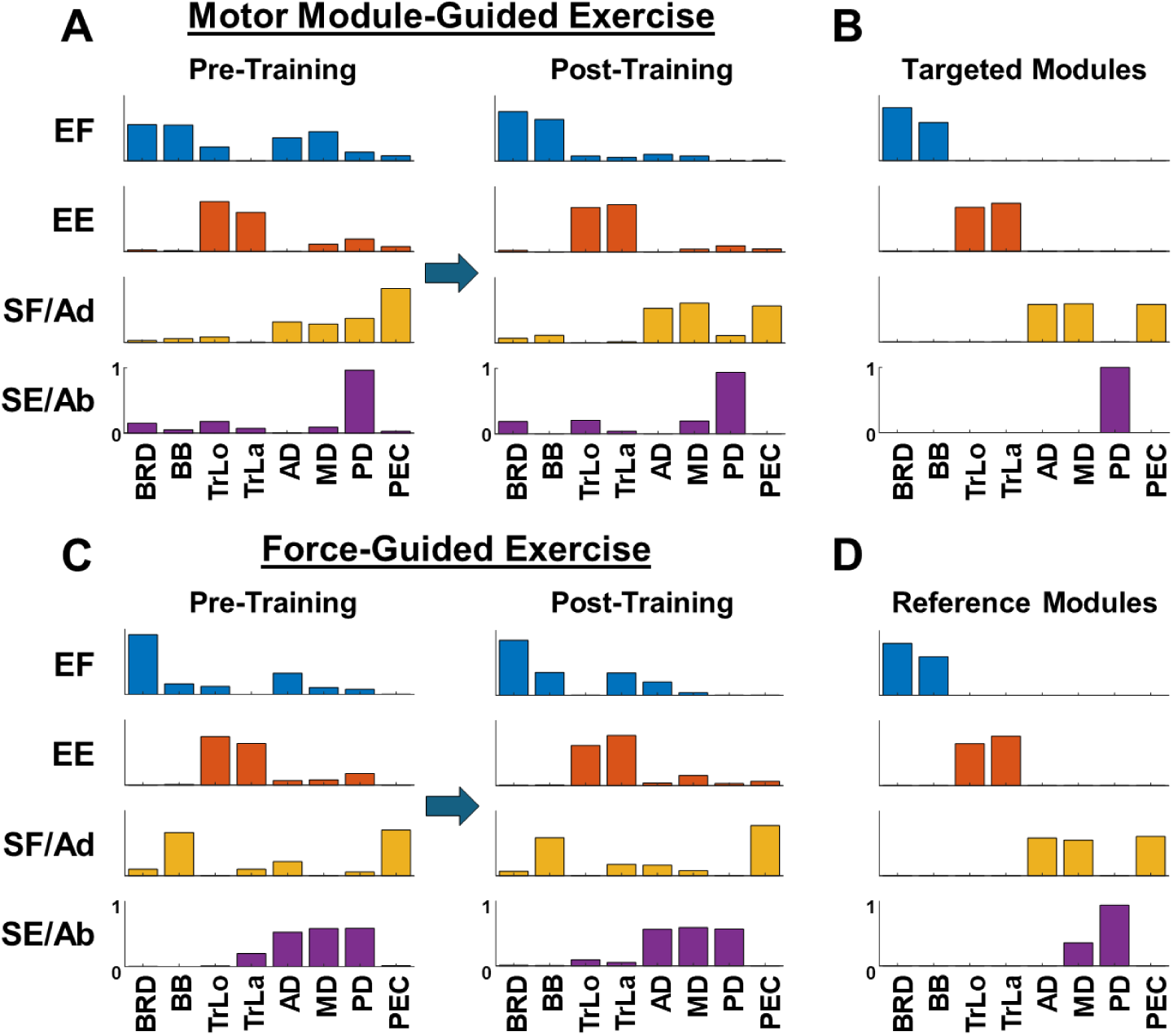
The motor module-guided feedback induced changes in the composition of motor modules in stroke, but force-guided feedback did not. **A,** Pre- and post-intermuscular coordination patterns after motor module-guided exercise in an illustrative participant (P1; stroke survivor with mild motor impairment). **B,** Customized targeted motor modules identified from P1’s non-paretic arm used for motor module-guided training. **C,** Pre- and post-intermuscular coordination after force-guided exercise in an illustrative participant (P7; stroke survivor with moderate motor impairment). **D,** Customized reference modules, identified from P7’s non-paretic arm, were not used as feedback during force-training. The four identified motor modules are shown (EF, elbow flexor; EE, elbow extensor; SF/Ad, shoulder flexor/adductor; and SE/Ab, shoulder extensor/abductor). The eight major UE muscles were: brachioradialis (BRD), biceps brachii (BB), triceps brachii, long (TrLo) and lateral (TrLa) heads, the three fibers of the deltoids (anterior (AD), middle (MD), and posterior (PD)), and the clavicular head of the pectoralis major (PEC).

In contrast, no participant in the force-guided exercise showed improvement in intermuscular coordination. For instance, the participant (P7) with moderate motor impairment (pre-training UE-FMA = 39 out of 66) in the force-guided group showed abnormal activation across multiple motor modules, including 1) atypical isolation of BRD in EF motor module; 2) abnormal co-activation of BB and PEC in the SF/Ad motor module, and altered co-activation of the three deltoid fibers in SE/Ab motor module (blue, orange and purple modules in **Fig. 6C**, respectively). These abnormal patterns did not change after force-guided exercise, nor did they resemble the reference modules identified from the non-paretic arm (**Fig. 6D**). The reference module targets in the force-guided group (**Fig. 6D**) are shown in the figure as a reference only. The reference modules were not used as a part of the actual training feedback, as the exercise of the active control group relied exclusively on force-guided feedback.

Notably, the results of the two exercises showed that improvement of the altered intermuscular coordination was associated with an increase in standardized clinical scores to assessment motor impairment. A positive correlation was found between the changes in UE-FMA and changes in the average of similarity indices of stroke-affected motor modules per participant (see Methods section “Calculation of Motor Module Similarity”; **Fig. 7**).

**Figure 7.**
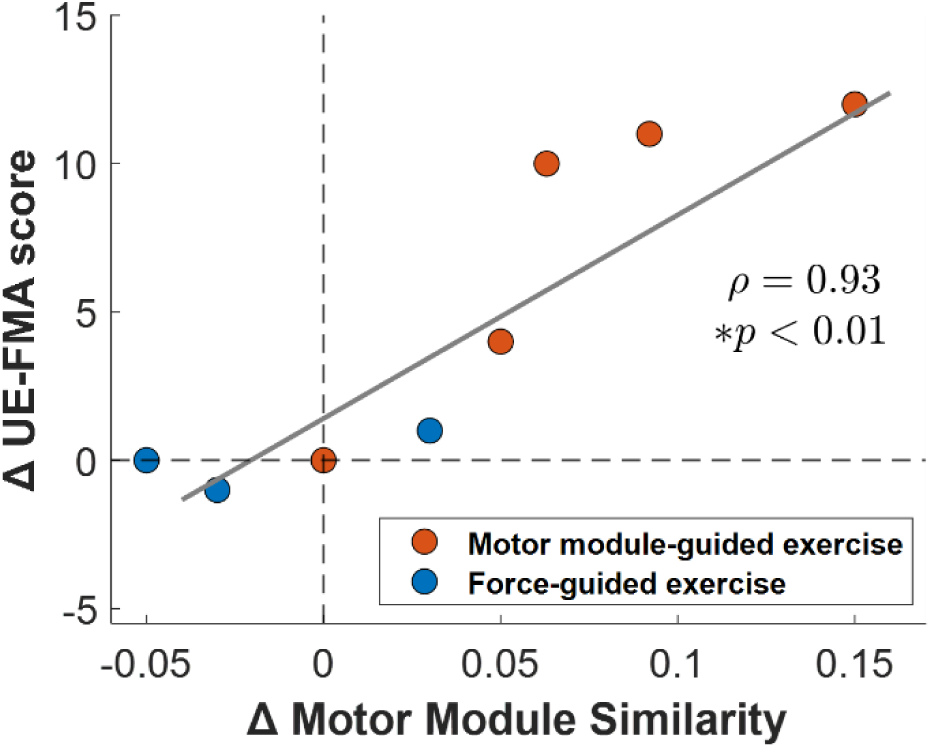
Correlation between changes in the upper extremity Fugl-Meyer Assessment (UE-FMA) and changes in the altered intermuscular coordination (motor module) similarity after six weeks of training for both exercises (Spearman correlation, rho = 0.93; *, p < 0.01). The regression line was drawn for visualization purposes only.

### Generalizability of Motor Learning

#### Intermuscular Coordination Underlying Dynamic Tasks

During 3D dynamic assessment, three of the four responders exhibited all four motor modules underlying the untrained dynamic reaching task with compositions statistically similar to those of age-range-matched healthy individuals after motor module-guided exercise (**Fig. 8**), suggesting potential transfer of the restored neuromuscular coordination from the trained isometric condition to the untrained dynamic reaching task. Notably, the non-responder also exhibited motor modules during the dynamic task that were similar to the healthy reference modules. In contrast, following force-guided training, only the mildly impaired participant exhibited all four motor modules similar to the healthy reference, whereas the moderately and severely impaired participants each retained two abnormal motor modules.

**Figure 8.**
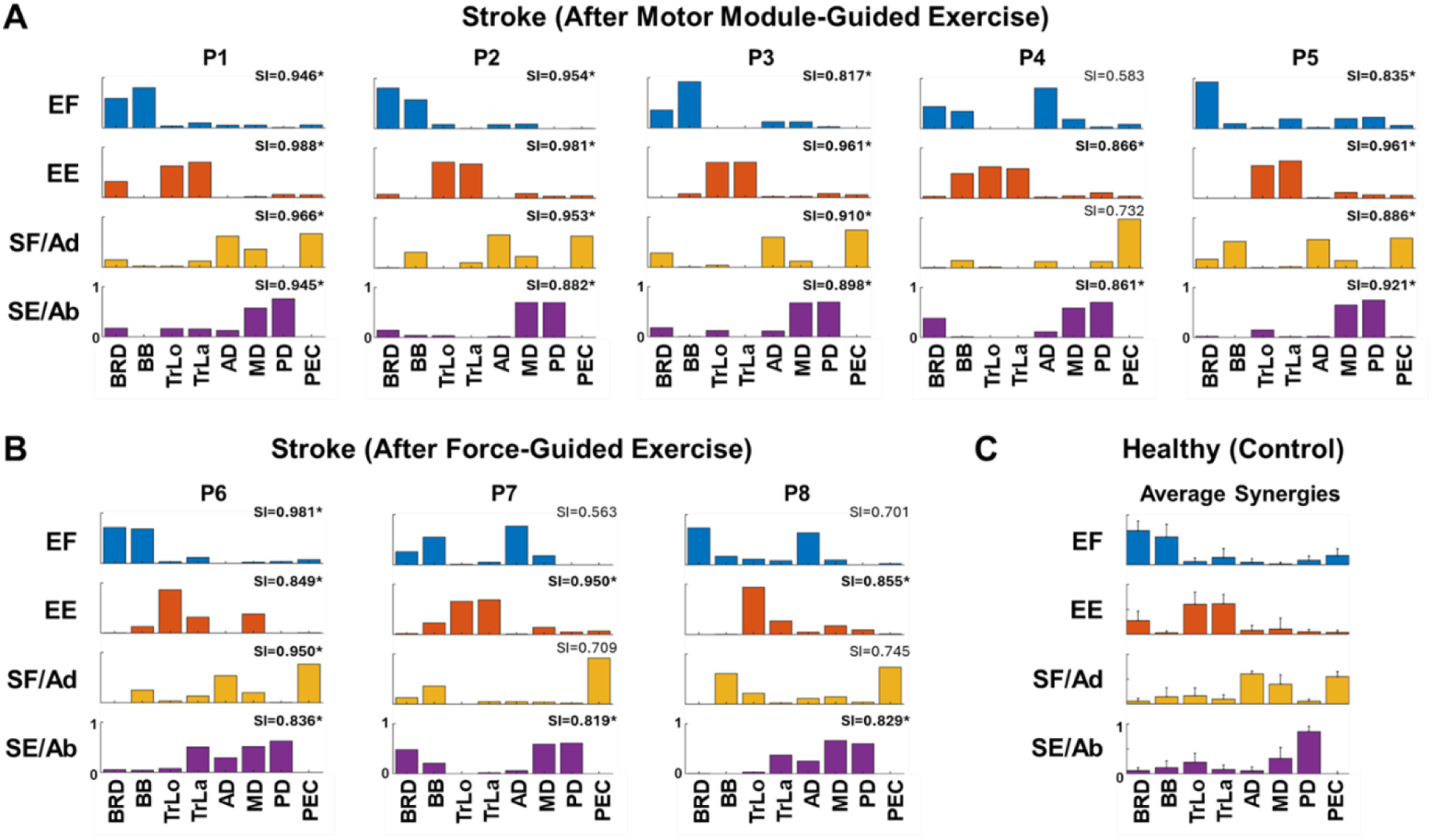
Composition of motor modules identified from dynamic motor tasks after the motor module-guided training (**A**), force-guided training in stroke (**B**), and reference (norm) motor modules (mean ± SD; n=8) of age-range-matched neurologically intact individuals from our previous study (**C**) [15]. The similarity indices (SI) shown next to each motor module indicate the similarity index between each stroke participant’s motor module and their corresponding norm modules. Values exceeding the dynamic threshold (0.814) were considered statistically similar (*, p < 0.05). EF, elbow flexor module; EE, elbow extensor module; SF/Ad, shoulder flexor/adductor module; SE/Ab, shoulder extensor/abductor module.

#### Changes in Kinematics During Dynamic Task Performance

The effect of isometric motor module-guided exercise extended beyond improvements in intermuscular coordination of the trained tasks to include enhancements in untrained, movement kinematics. **Figure 9A** shows that both exercise groups exhibited an increase in kinematic synergy similarity—representing inter-joint coordination during dynamic tasks (point-to-point reaching and drinking)—relative to healthy controls. However, improvements were generally greater after motor module-guided exercise, among participants with mild and moderate impairment. Motor module-guided exercise also facilitated improvements in pairwise joint angle-to-angle correlations during point-to-point reaching (**Fig. 9B**). The shifts in correlation coefficients (r) toward more healthy patterns in the motor module-guided group were comparable to, or greater than, those observed in the force-guided exercise group. In addition, the non-responder (P5) in the motor module-guided group showed no improvement in inter-joint coordination, aligning with their lack of improvement in motor impairment (**Fig. 9A and 9B**). Since the participants with severe impairment could not complete the unsupported dynamic tasks, ARoM of shoulder and elbow flexion was assessed instead of kinematic synergies in three participants with severe impairment (P2, P4, and P8). The motor module-guided exercise group demonstrated greater improvements in ARoM compared to force-guided exercise (**Fig. 9C**). We observed participant-specific recovery patterns within the motor module-guided group. For example, participant P2 exhibited a substantial increase in shoulder flexion while P4 showed primary improvements in elbow flexion. In contrast, participants with severe impairment in the force-guided group (P8) showed minimal to no change in ARoM for either joint.

**Figure 9.**
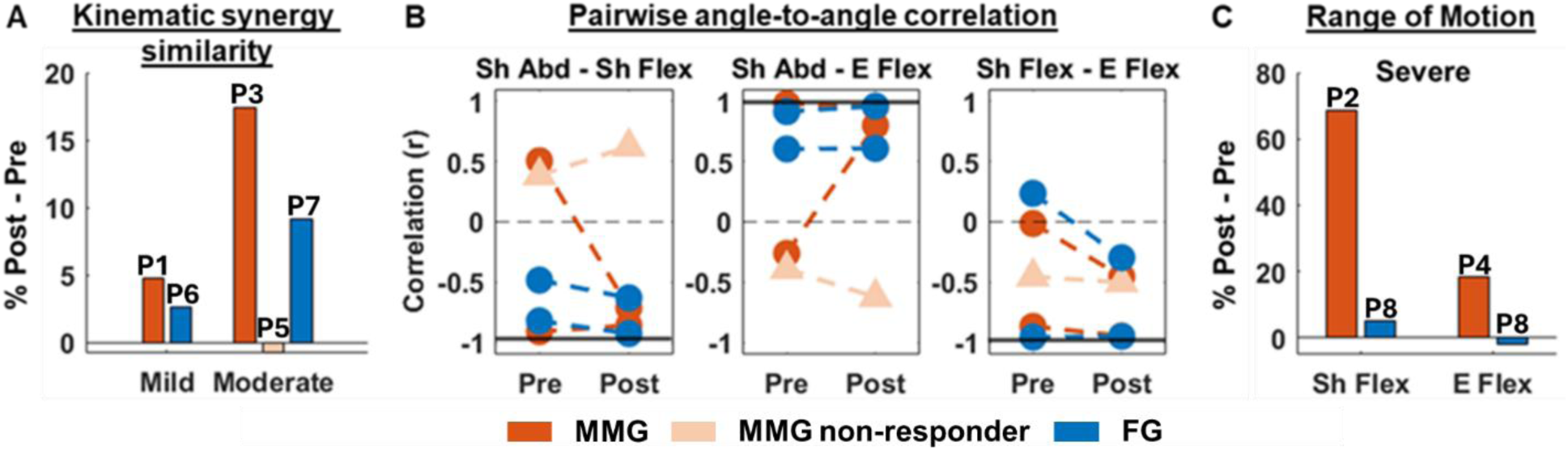
The effects of isometric motor module-guided and force-guided human-machine interaction on untrained, dynamic motor task performance in chronic stroke. A, Improvement of inter-joint coordination, reflected in kinematic synergy similarity. B, Joint angle-to-angle correlation. The black solid lines indicate a representative value from a healthy individual. C, The range of motion during shoulder and elbow flexion of the participants who could not complete the dynamic movements in A.

#### Spontaneous Comments about Improvements in Activities of Daily Life

In addition to the quantitative improvements, the four responders in the motor module-guided training group shared spontaneous, qualitative changes in their daily function (**Table 2**), especially on muscle strength, muscle tone, and range of motion. The participant with mild motor impairment (P1) expressed that she felt her arm was "stronger than before" and reported a major personal milestone: being able to perform a push-up for the first time since her stroke. The participant with severe motor impairment (P2) emphasized the sense of relaxation he experienced in his left arm (the paretic one) after training, which allowed him to sleep in his preferred sleep posture (sleeping on his left side) again. The participant with moderate motor impairment (P3) described improved range of motion, enabling him to use both hands to wash his hair. Finally, the other participant with severe motor impairment (P4) reported improved ability to extend his arm and maintain it in a lower position during walking.

**Table 2.** Spontaneous comments were provided by participants over the course of motor module-guided exercise.

|  | **Positive comments provided by participants over the course of motor-module-guided exercise** |
| --- | --- |
| **P1 - <u>Mild</u>** impairment<br>(right arm affected) | "I am right-handed, but after my stroke, I learnt to use my left hand to eat. Now, I eat with my right hand again."<br>**"I feel my arm is stronger.** I try to exercise at home, and I could finally do a push-up since my stroke." |
| **P2 - <u>Severe</u>** impairment<br>(left arm affected) | **"I feel my arm more relaxed."**<br>"I couldn't sleep on my left side due to discomfort before. Now, it is easier to change positions in the bed, and I can comfortably sleep on both sides." |
| **P3 - <u>Moderate</u>** impairment<br>(right arm affected) | "I was not able to wash my hair with my right arm after my accident, but yesterday, I used both hands."<br>"I dropped my car keys. I tried to pick them up with my right hand. I did it! **I was not able to reach things on the floor before.**" |
| **P4 - <u>Severe</u>** impairment<br>(right arm affected) | **"I can move my arm down better."**<br>"While walking holding hands with my wife, she tried to lower my arm, but I used to pull her arm up. Now I can keep my arm lower when we walk." |

P5 – **Moderate** impairment: Participant did not report any spontaneous comment during or after motor-module-guided exercise.

## DISCUSSION

The aim of this pilot study was to develop and evaluate whether a novel neuromuscular coordination-guided intervention could promote recovery of abnormal intermuscular coordination and reduce motor impairment in chronic stroke survivors with different levels of motor impairment. A force-guided training condition served as an active control exercise. Traditionally, the concept of motor modules has primarily been used to characterize neuromuscular coordination deficits following stroke, with limited translation into a therapeutic framework. In this study, motor module analysis was translated into a novel rehabilitation strategy design through the implementation of a personalized, motor module-guided training protocol. Three principal findings emerged. First, abnormal intermuscular coordination of the upper extremity was amenable to improvement through this innovative human-machine interaction in chronic stroke survivors. Second, four out of five stroke survivors who participated in the motor module-guided training showed a significant improvement in clinical scores that indicated reduction in motor impairment. Third, the results suggest potential transfer of training-related benefits from the practiced isometric task to untrained dynamic motor tasks, supporting the premise that neuromuscular-coordination-based rehabilitation may generalize positive effects beyond the specific training context.

### Effects of Motor Module-Guided Training on Neuromuscular Coordination and Motor Impairment

A well-documented consequence following stroke is the development of abnormal stereotypical movement patterns, impaired intermuscular coordination, and loss of independent joint control, whereby voluntary activation of muscles at one joint elicits involuntary activation at adjacent joints[5], [11], [58], [59]. One common example of this phenomenon is the clinical flexion synergy, in which increasing shoulder abductor activation drives progressive, involuntary activation of elbow, wrist, and finger flexion [22]. Accumulating evidence indicates that these stereotypical multi-joint coupling patterns might involve multifactorial changes, including the corticospinal tract damage, upregulation of brainstem pathways (particularly the medial reticulospinal tract, and potentially the vestibulospinal tract), and maladaptive spinal network plasticity [22], [25], [58]. Following corticospinal tract damage, increased reliance on the reticulospinal projections — which are anatomically diffuse and cannot produce discrete, joint-specific movements — likely results in atypical co-activation across multiple joints rather than selective single-joint control [5]. These findings provide a clear rationale for developing novel rehabilitation that aims at directly targeting abnormal neuromuscular coordination, rather than impaired movements alone, to reduce motor impairment and improve individual joint control after stroke.

Previous studies have documented alterations in neuromuscular coordination characteristics (e.g., motor module composition) after stroke across varying biomechanical conditions [11]. For instance, under isometric force generation conditions, stroke induces abnormal coactivation of the three deltoid fibers within the shoulder abductor/extensor motor module, with the prevalence of this alteration increasing as motor impairment severity increases [13]. Furthermore, abnormal coactivation between elbow flexors and shoulder adductors/flexors has been found, particularly in severely impaired stroke survivors, not only under isometric force generation conditions but also during dynamic reaching [15]. Another study investigating functional dynamic conditions (i.e., hand-to-mouth and reaching movements) found that merging was the most common abnormal motor module pattern, particularly in moderately impaired stroke survivors [20]. Collectively, these findings underscore the clinical need for rehabilitation strategies capable of directly addressing the reorganization of multiple synergistic muscle groups after stroke.

To our knowledge, the current study is the first to provide evidence that a non-invasive, closed-loop motor module-guided human-machine interaction paradigm can retrain intermuscular coordination patterns toward healthy ones. This finding is notable because previous EMG-guided rehabilitation paradigms have primarily focused on biofeedback from individual muscles or a small number of muscle pairs to improve selective muscle activation and reduce abnormal co-activation [34], [35], [36], [37], [38], [39], [60]. Although these approaches have demonstrated improvements in task performance and muscle activation, direct evidence of reshaping multi-muscle motor module composition toward healthy patterns has remained limited [11]. In addition, a major limitation of traditional EMG-guided approaches is that reported improvements have primarily been demonstrated in the short term, while evidence supporting long-term (e.g., more than a month) benefit remains limited [33]. Recently, motor module-guided rehabilitation strategies have emerged. These approaches can be broadly divided into two categories: interventions that use motor modules to generate stimulation patterns passively delivered to stroke survivors, such as synergy-based functional electrical stimulation (FES) [41], and interventions that provide motor modules as real-time biofeedback, requiring participants to actively modulate their neuromuscular coordination. Synergy-based FES studies have reported shifts toward healthier motor module composition together with functional improvements [40], [41], [42]. However, synergy-based FES relies on externally generated coordination patterns, with participants having limited control over the formation of the stimulation-induced motor modules. Thus, the extent to which the observed shifts in motor module composition reflect transient biomechanical entrainment or endogenous neural reorganization of the motor control architecture remains unclear. In addition, further investigation is needed to determine the long-term retention and transferability of these adaptations to untrained tasks. In contrast, our motor module-guided biofeedback paradigm required participants to volitionally activate and enhance control of each motor module throughout training. By requiring participants to actively modulate their neuromuscular coordination, this approach might directly engage the CNS in reorganizing its modular control architecture, potentially promoting endogenous neuroplastic changes that could be more durable and transferable than those achieved through externally imposed stimulation patterns. The present findings therefore support the feasibility of using motor module-guided biofeedback to directly retrain impaired neuromuscular coordination after stroke.

The motor module–guided training paradigm developed in this study was specifically designed to train desired intermuscular coordination and enhance individual joint control to reduce motor impairment. Accordingly, four motor module targets were designed to train stroke survivors to independently control synergistic muscle groups (i.e., elbow flexors, elbow extensors, shoulder extensors/adductors, and shoulder flexors/abductors) one at a time and suppress the activation of three others. We designed the targeted motor modules based on patterns observed in neurologically intact individuals [13], [15], [31], [45] and customized them using each participant’s non-paretic arm. By requiring participants to selectively activate a single synergistic muscle group (e.g., elbow flexors) while simultaneously minimizing activation of all other muscle groups — including the antagonist at the same joint (e.g., elbow extensors) and muscles acting at adjacent joints (e.g., shoulder muscles) — the training inherently demanded independent control of joint-specific muscle groups (**Fig. 2**). Successful target matching therefore required participants to decouple the stereotypical multi-joint co-activation patterns that characterize post-stroke motor impairment. Thus, the increase in the number of matched targets observed after six weeks of motor module– guided training (**Fig. 4**) suggests that participants progressively enhanced the ability to isolate and independently recruit the four targeted synergistic muscle groups, reflecting an improvement in joint-specific motor control. This interpretation is consistent with the paradigm design, in which co-activation of antagonistic motor modules within the same joint canceled cursor movement (**Fig. 2C**), and co-activation across joints moved the cursor away from the intended target (**Fig. 2D**), thereby reinforcing selective, individualized joint control throughout the training process. Notably, the specific motor module targets that proved most challenging varied across participants, with no single target consistently emerging as the most difficult in the last week of training (**Fig. 4**). This variability in target difficulty across post-stroke participants likely reflects individual differences in the pattern of neuromuscular coordination impairment after stroke, as well as potential variability in performance across stroke survivors. This observation suggests that training protocols designed around each participant’s own non-paretic arm coordination may more effectively address their specific deficits than a one-size-fits-all rehabilitation approach.

Alteration in motor module composition after stroke has been consistently associated with the severity of motor impairment [11], which decreases the ability to perform activities of daily living. For instance, the severity of motor impairment has been shown to correlate negatively with the similarity of post-stroke muscle synergies relative to those of neurologically intact individuals [11], and the prevalence of abnormal motor module patterns tends to increase with greater impairment severity [13], [45]. In the present study, improvements in neuromuscular coordination were accompanied by increases in the UE-FMA score (**Fig. 8**), suggesting that shifting stroke-induced intermuscular coordination toward patterns observed in healthy individuals may contribute to reductions in motor impairment. Four out of the five participants exceeded the minimal clinically important difference (≥ 4 points [57]) after the motor module-guided training, with an average improvement of 9.25 points. Notably, these positive effects were typically maintained at 1- and 3-month follow-ups, indicating potential long-term retention of motor gains associated with the motor module-guided protocol (**Fig. 5**). Although confirmation in a larger study is required, these findings provide preliminary evidence that motor module-guided biofeedback may promote longer-lasting benefits than those currently reported for conventional EMG-guided rehabilitation approaches. In comparison, no participant in the active control group showed clinically meaningful improvement in UE-FMA following the six-week force-guided exercise, suggesting that force generation alone may be insufficient to drive comparable reductions in motor impairment. Interestingly, following motor module-guided training, the ARAT improvements were primarily observed in participants with moderate levels of motor impairment (**Supplementary Fig. 1**), suggesting that individuals within this impairment range may have sufficient residual motor capacity to benefit from neuromuscular coordination retraining while still exhibiting room for functional improvement. In contrast, the absence of ARAT changes in participants with milder or more severe impairment may be partially related to potential ceiling and floor effects [61], [62], respectively. However, given the limited sample size, future studies with larger cohorts are required to determine whether baseline impairment severity influences responsiveness to motor module-guided training and how restoration of neuromuscular coordination contributes to improvements in functional upper extremity movements. Overall, the converging evidence from previous studies and the present results supports the premise that motor module analysis can serve as a valuable tool for both assessing neuromuscular coordination and informing the design of novel rehabilitation strategies to reduce motor impairment after stroke.

### Motor Learning Transferability

One of the main goals of stroke rehabilitation is to induce favorable neuroplastic changes that enhance motor function; consequently, promoting the transferability of training gains beyond the practiced task is fundamental to effective intervention. Nevertheless, previous studies have shown that task-specific training improves performance in practiced tasks, whereas the improvement in the untrained task was not as significant [63], [64]. In addition, improvements in motor capacity do not automatically translate into improved functional movement in activities of daily living [65]. For these reasons, targeting fundamental bases of neuromuscular control (e.g., motor module characteristics) instead of focusing on the resultant symptoms (e.g., impaired functional movement) may facilitate the transfer of benefits from trained tasks to untrained tasks. In this pilot study, evidence of such transferability was observed in the motor module-guided training group. First, despite both exercises being performed under isometric conditions, the motor-module-guided training group showed greater improvements in kinematic synergy similarity during dynamic movements in participants with mild-to-moderate impairment (**Fig. 8A**). In addition, its significant association with motor module similarity suggests that improvements in intermuscular coordination under isometric conditions can positively influence joint coordination during dynamic movements (**Supplementary Fig. 4A**). The concurrent validity of the kinematic synergy similarity measure was further supported by a significant, monotonic relationship with the clinical score (**Supplementary Fig. 4B**). Second, pairwise angle-to-angle coordination during point-to-point reaching revealed similar or greater improvement in mildly and moderately impaired participants after motor module-guided exercise compared with force-guided exercise (**Fig. 8B**). This may suggest that motor module-guided exercise helps achieve targeted reaching movement through promoting true recovery in terms of joint coordination, rather than through compensatory movements. Third, both severely impaired participants after motor module-guided exercise showed much greater improvement in range of motion P2 in shoulder flexion and P4 in elbow flexion compared with the severely impaired participant (P8) in the force-guided exercise (**Fig. 8C**). These findings suggest that while the motor-module guided exercise is effective for severe cases, the specific joint recovery may vary by individual. Fourth, following motor module-guided exercise, intermuscular coordination patterns identified from a 3-D dynamic point-to-point reaching task resembled those identified in healthy individuals (**Fig. 9A**), suggesting that improvements in intermuscular coordination achieved under isometric conditions may generalize to dynamic tasks, even in participants with moderate and severe impairment. This finding is compatible with our previous studies, which showed that the composition of muscle synergies is largely conserved across isometric and dynamic conditions in both neurologically intact individuals [31] and chronic stroke survivors [15]. Lastly, the spontaneous comments of participants suggest that motor module-guided training may translate into meaningful functional gains not fully captured by standard clinical measures (**Table 2**). Improvements in perceived strength, relaxation, and bilateral coordination indicate potential benefits in both motor control and quality of life. Additionally, regaining the ability to perform everyday tasks highlights the potential value of targeting intermuscular coordination to support functional recovery after stroke. Overall, these results support the premise that transferability of training effects may be achieved when targeting underlying pathophysiological mechanisms (i.e., abnormal intermuscular coordination) rather than task-specific deficits.

### Biomechanical Considerations and Future Direction

An important aspect of any rehabilitation strategy is the selection of the biomechanical context in which assessments and interventions are delivered. In this study, the motor module-guided paradigm was implemented under isometric conditions for several reasons. First, stroke survivors with severe impairment often are unable to perform goal-directed dynamic reaching movements; however, they still could generate isometric forces in some 3D directions [13], [15], showing that isometric exercises are more accessible than movement conditions. Second, stroke induces alterations in motor module composition underlying isometric conditions [13], [15], [45], suggesting that this biomechanical context could provide a relevant framework for targeting abnormal neuromuscular coordination. Third, our previous study showed that chronic stroke survivors, even with severe motor impairment, were able to increase the repertoire of intermuscular coordination patterns after six weeks of isometric exercise, showing evidence that motor module composition can be modulated through this biomechanical condition [66]. Fourth, in our previous assessment studies, we showed evidence of neuromuscular coordination patterns shared between isometric force generation and dynamic reaching conditions in healthy individuals [31] and chronic stroke survivors [15]. These studies suggest that modulating motor module composition in a training task (e.g., isometric) could transfer the benefits to untrained tasks (e.g., dynamic reaching).

Improvements in motor impairment associated with enhanced neuromuscular coordination after the motor module-guided exercise under isometric conditions raise an important question: Do the changes in motor module composition reflect a learned ability to individually activate synergistic groups of muscles, or are they primarily the result of repetitive isometric force production in specific directions (i.e., motor module preferred directions [MMPD])? Accordingly, a force-guided exercise was designed as active control training to account for this potential confounding factor in our study. Both interventions (i.e., motor module-guided and force-guided exercises) shared critical factors, such as four training targets, 18 training sessions within six weeks, and arm position during training. However, neuromuscular coordination training and force training differ in two main characteristics: 1) the control signals (EMG from eight UE muscles and 3D isometric forces measured at the hand, respectively); and 2) the four target setups (synergistic muscle targets and force targets). Thus, the experimental design was intended to distinguish the effects of neuromuscular coordination training from those of repeated force production without muscle coordination feedback. In the control group, participants showed an increase in the number of matched targets after six weeks of force-guided training (**Fig. 4**), demonstrating improvement in training task performance, particularly in the ability to accurately generate and stabilize isometric force magnitude in the four directions (i.e., MMPDs). Notably, only the participants in the control group exhibited a reduction in the number of peaks during the 3D unbiased isometric assessment (**Supplementary Fig. 5**), suggesting enhanced force control following force-guided training, but not observed in the motor module-guided group. Additionally, following the force-guided training, two out of three participants improved their ARAT score by 3 points (**Supplementary Fig. 1**). However, in the control group, no participant showed improvement in intermuscular coordination patterns or clinically meaningful improvement in UE-FMA. Overall, these findings suggest that although force-guided training may contribute to improvements in force control, task performance and upper extremity function, practicing force generation in the four MMPDs alone may be insufficient to modulate motor module composition and reduce motor impairment.

Within the broader framework of EMG biofeedback in neurorehabilitation, interventions can be conceptually divided into motor module-guided approaches — which target multi-muscle coordination patterns — and non-motor-module methods, which typically provide biofeedback based on the amplitude of individual muscles or a small number of muscle pairs [34], [35], [67]. Viewed through this framework, a potentially more closely matched active comparator for the motor module-guided training would be an EMG amplitude-guided rehabilitation paradigm. In such a design, participants would receive biofeedback based on the EMG amplitude of the muscles within a single targeted synergistic group (e.g., elbow flexors), requiring activation of that group to match a target, but without providing information about the activity of muscles belonging to other motor modules. This approach would offer several advantages over the force-guided control used in the present study. Most notably, both the motor module-guided and the EMG amplitude-guided paradigms would share the same control signal type (i.e., EMG), the same sensor placement, and a comparable human-machine interaction experience, thereby minimizing confounding differences inherent to comparing fundamentally different task modalities (i.e., EMG-based versus force-based control). The primary limitation of the force-guided control is that the two interventions are inherently different tasks — one driven by neuromuscular signals and the other by mechanical output — making it difficult to determine whether the observed benefits of the motor module-guided training arise specifically from the coordination-based feedback or from broader differences in task demands and sensorimotor engagement. An EMG amplitude-guided comparator might more precisely isolate the contribution of multi-muscle coordination feedback by controlling for the use of EMG-based biofeedback itself. However, given the pilot nature of the present study, the force-guided control was selected to investigate whether targeting intermuscular coordination patterns produces benefits beyond those attributable to repetitive isometric force generation without learning desired muscle coordination. Future adequately powered studies can incorporate an EMG amplitude-guided comparator to more precisely delineate the added value of neuromuscular coordination-guided feedback.

We observed that one participant (P5) was a non-responder whose UE-FMA score did not improve after the motor module-guided exercise in this pilot study. Interestingly, this participant exhibited pre-training intermuscular coordination patterns in the paretic arm that closely resembled those of healthy individuals (**Supplementary Fig. 3**). While the sample size of this study is small, this non-responder case provides preliminary insight into who may benefit the most from this intervention. Specifically, stroke survivors with relatively preserved neuromuscular coordination at baseline may have limited capacity for further improvement through this motor module-guided paradigm. Although these observations are exploratory and not part of the primary findings, they suggest that baseline neuromuscular organization may influence responsiveness to this type of exercise. This consideration may inform future refinement of inclusion and exclusion criteria and support more targeted application of motor module-guided rehabilitation strategies.

Finally, the present study also suggests that further refinement of the motor module-guided training protocol may be beneficial, particularly for stroke survivors with severe motor impairment. In one participant (P2; baseline FMA-UE = 6 out of 66), the initial training difficulty exceeded the participant’s ability to successfully interact with the paradigm, requiring adjustment of the training parameters and an extended training period. Following these modifications, continued improvements in motor impairment were observed, suggesting that individuals with more severe impairment may benefit from personalized progression of training difficulty and longer intervention durations. Indeed, systematically matching task difficulty to patient ability has been shown to improve upper extremity motor function after stroke, reinforcing the importance of calibrating training demands to the individual’s current capacity [68]. Future studies are needed to systematically optimize motor module-guided training parameters and progression strategies according to individual patient characteristics (e.g., baseline motor impairment and chronicity of stroke).

### Limitations

There are several limitations in this pilot study. First, the number of participants is relatively small; consequently, while our preliminary results showed the feasibility of restoring intermuscular coordination patterns to reduce motor impairment, a larger sample is necessary to validate these results. In addition, no consistent changes in the activation profiles across participants were found, likely due to the small sample size and the differences in the level of motor impairment. Second, the training protocols did not incorporate gamification or adaptive difficulty. Although improvement in motor impairment was observed, the lack of motivational features may limit engagement and adherence. Future studies could gamify and adjust the level of difficulty to increase participant engagement during exercise performance, particularly in less-supervised settings such as home-based rehabilitation. Third, we showed evidence that in the motor module-guided group, post-training intermuscular coordination underlying dynamic conditions resembles the one identified in healthy individuals. However, pre-training motor modules during dynamic conditions from the same individuals were not collected in this pilot study, limiting the ability to quantify the changes in neuromuscular coordination during dynamic reaching induced by the motor module-guided isometric exercise. Fourth, potential transferability to other UE segments was not tested, even though it is feasible (**Fig. 5C)**. For instance, future studies could investigate how modulating motor modules in proximal joints (e.g., shoulder and elbow) could benefit more distal joints (e.g., wrist and fingers) [69], [70] by assessing motor function (e.g., ARAT or Wolf Motor Functional Test) or identifying hand neuromuscular coordination across various biomechanical conditions.

### Implications for Neurorehabilitation

This pilot study provides important implications for neurorehabilitation. First, retraining desired neuromuscular coordination patterns through a non-invasive human-machine interaction is feasible in chronic stroke. Second, improvements in motor module composition are correlated with a reduction of motor impairment, providing a new framework for rehabilitation. Third, isometric rehabilitation interventions could be used as an alternative effective rehabilitation strategy to improve motor function, even in severely impaired stroke survivors. Fourth, our preliminary evidence of transfer across biomechanical conditions suggests that improvements in intermuscular coordination achieved through isometric exercise may generalize to functional dynamic reaching. Future directions of this work include: 1) increasing the sample size to conclude it as a full study; 2) translating this rehabilitation protocol to home therapy; 3) investigating whether a similar intervention could improve neuromuscular coordination in the lower extremity or hand.

## Data Availability

Availability of data and materials
The data used in this study may be made available by the corresponding author upon a reasonable request.

## Abbreviations

UE: Upper extremity
KULSIS: The KAIST Upper Limb Synergy Investigation System
EF: Elbow Flexor
EE: Elbow Extensor
SF/Ad: Shoulder Flexor/Adductor
SE/Ab: Shoulder Extensor/Abductor
MMPD: Motor module preferred direction

## Declarations

### Availability of data and materials

The data used in this study may be made available by the corresponding author upon a reasonable request.

### Contributions

MPJ, GS, YNGH, and MH completed the data collection. MPJ, YNGH, MH, and SV performed the data analysis. MPJ, GS, YNGH, MH & SV contributed to manuscript writing. MPJ, GS, YNGH & JR originally developed the study design. YZ, H-SP, SL, and JR further refined the study design, contributed to the interpretation of the results, and helped write the manuscript. MPJ and GS developed training software. All authors read and approved the final manuscript.

## Acknowledgments

The authors would like to thank Komal Kukkar for his contributions to clinical assessments.

## Ethics declarations

### Ethics approval and consent to participate

The research study was approved by the University of Houston Institutional Review Board (STUDY000013333), and each participant gave written informed consent before the beginning of the study.

### Consent for publication

Not applicable.

