## Supplementary Information for "Motor Module-Guided Human-Machine Interaction Promotes Improvement of Neuromuscular Coordination and Reduction in Motor Impairment in Chronic Stroke: A Pilot Study"

### Action Research Arm Test (ARAT)

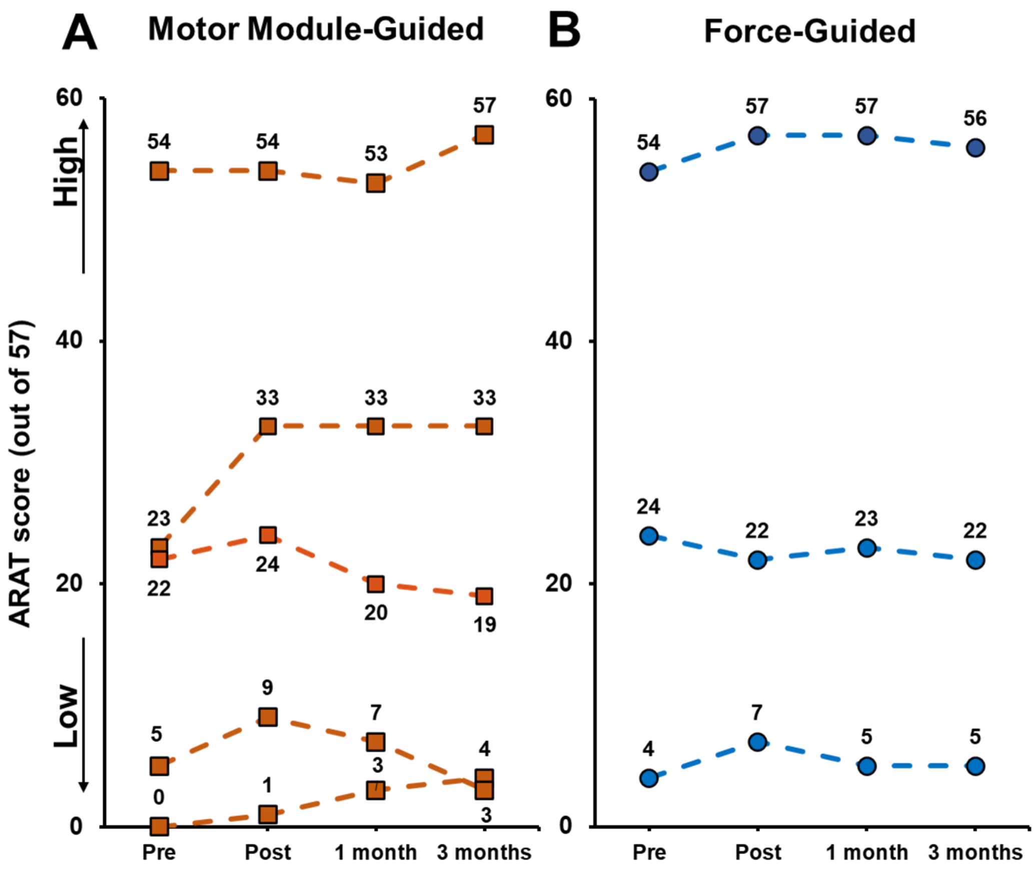


C

| | Motor Module-Guided Exercise | | | | | | | | | | Force-Guided Exercise | | | | | |
|---|---|---|---|---|---|---|---|---|---|---|---|---|---|---|---|---|
| | P1 | | P2 | | P3 | | P4 | | P5 | | P6 | | P7 | | P8 | |
| | pre | post | pre | post | pre | post | pre | post | pre | post | pre | post | pre | post | pre | post |
| Grasp (max = 18) | 18 | 18 | 0 | 0 | 12 | 12 | 0 | 2 | 9 | 9 | 18 | 18 | 9 | 10 | 0 | 0 |
| Grip (max = 12) | 12 | 12 | 0 | 0 | 5 | 7 | 2 | 4 | 5 | 6 | 10 | 12 | 6 | 3 | 0 | 4 |
| Pinch (max = 18) | 15 | 15 | 0 | 0 | 2 | 9 | 0 | 0 | 2 | 3 | 17 | 18 | 3 | 3 | 1 | 0 |
| Gross Movement (max = 9) | 9 | 9 | 0 | 0 | 4 | 5 | 3 | 3 | 6 | 6 | 9 | 9 | 6 | 6 | 3 | 3 |
| Total (max = 57) | **54** | **54** | **0** | **0** | **23** | **33** | **5** | **9** | **22** | **24** | **54** | **57** | **24** | **22** | **4** | **7** |
| **Δ Total (Post-Pre)** | **0** | | **0** | | **10** | | **4** | | **2** | | **3** | | **-2** | | **3** | |

***Supplementary Figure 1.*** *Action Research Arm Test (ARAT) pre- and post- training, and 1-month and 3-months after the last training session for both motor module-guided (**A**) and force-guided (**B**) exercises. **C,** Table to detail the ARAT values in both exercises. Pre, pre-training; post, post-training.*

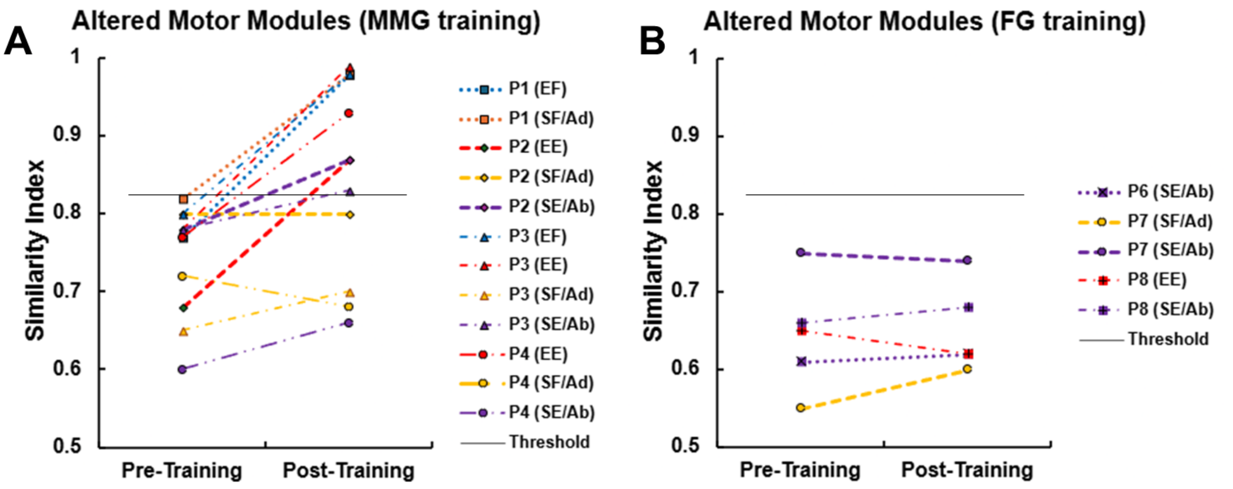


***Supplementary Figure 2.*** *Similarity indices between each participant's pre- or post-training synergy and its corresponding customized targeted synergies. The increase in the similarity indicates the improvement of intermuscular coordination after the training. Motor module-guided exercise (A) and force-guided exercise (B). EF, Elbow Flexor; SF/Ad, Shoulder Flexor/Adductor; EE, Elbow Extensor; SE/Ab, Shoulder Extensor/Abductor. The thin black solid line represents the similarity threshold (0.822).*

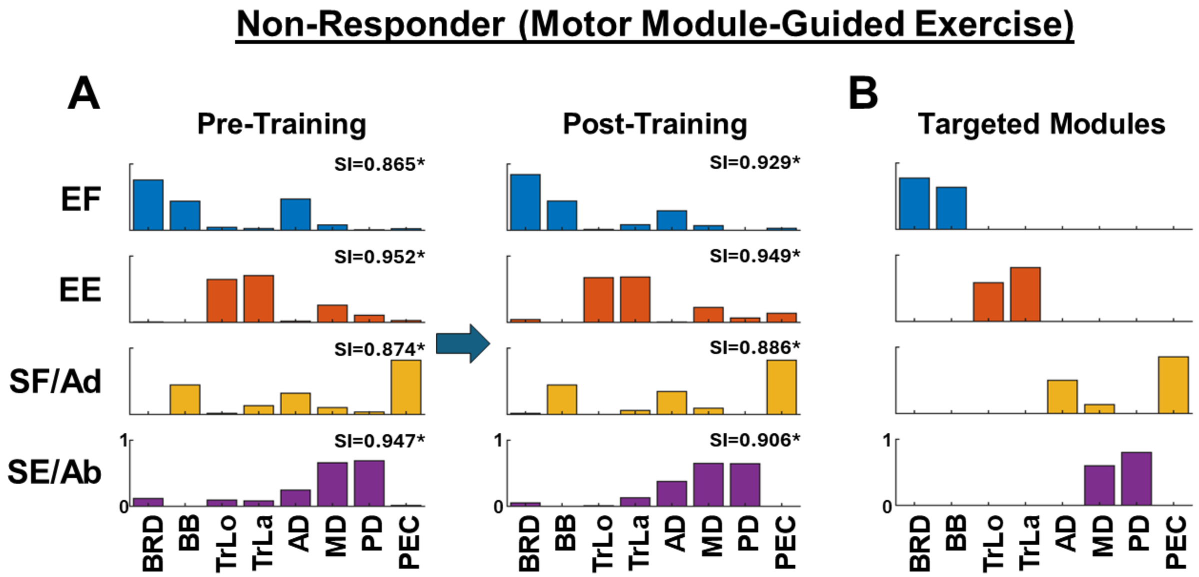


***Supplementary Figure 3.*** *Motor module changes for the non-responder participant (P5; stroke survivor with moderate motor impairment). A) Pre- and post-intermuscular coordination patterns following motor module-guided exercise. B) Customized targeted motor modules identified from P5's non-paretic arm used for motor module-guided training. The four identified motor modules are shown (EF, elbow flexor; EE, elbow extensor; SF/Ad, shoulder flexor/adductor; and SE/Ab, shoulder extensor/abductor). The eight major UE muscles were: brachioradialis (BRD), biceps brachii (BB), triceps brachii, long (TrLo) and lateral (TrLa) heads, the three fibers of the deltoids (anterior (AD), middle (MD), and posterior (PD)), and the clavicular head of the pectoralis major (PEC). The SI values indicate the similarity indices between each motor module from the paretic arm and its corresponding targeted motor module customized from the non-paretic arm. Note that the SI values were already statistically similar (threshold = 0.822) before motor-module training, suggesting that the paretic arm exhibited neuromuscular coordination patterns comparable to those of the non-paretic arm prior to the intervention.*

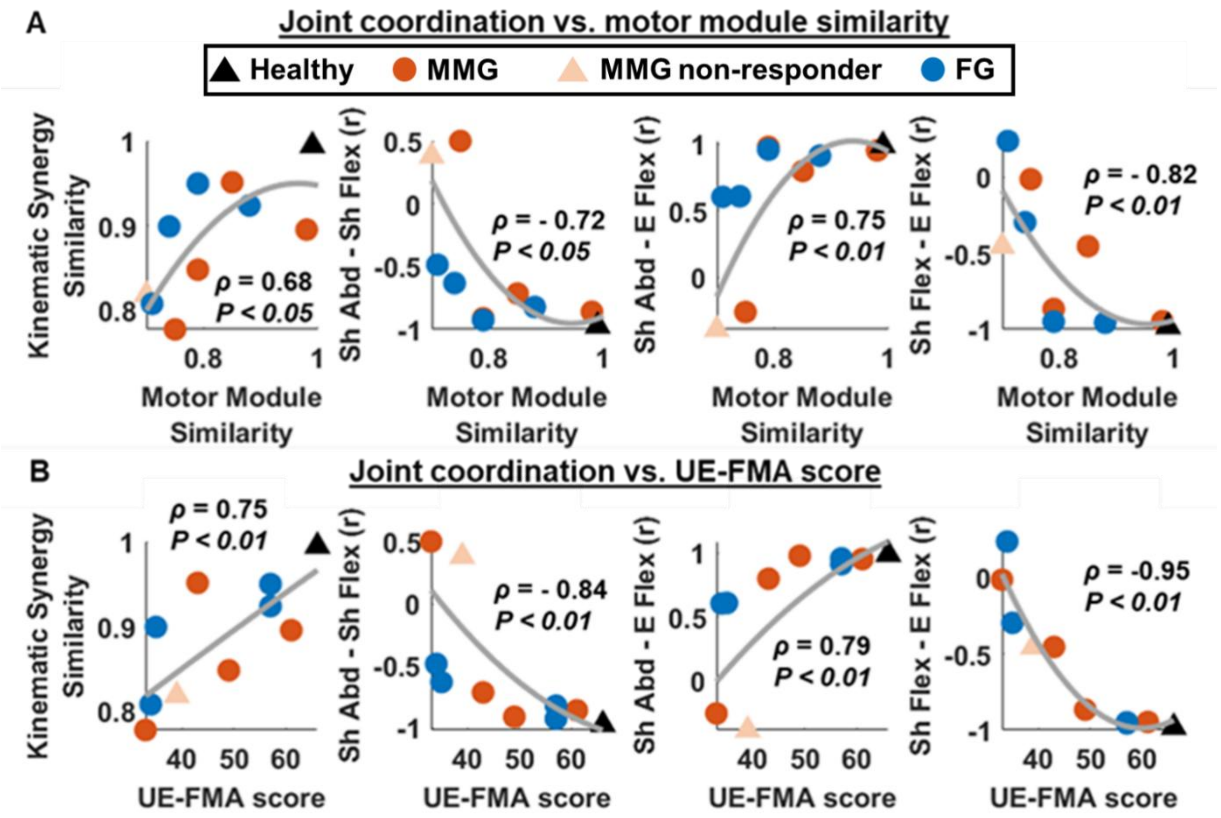


***Supplementary Figure 4.*** *Correlation between inter-joint coordination (i.e., kinematic synergy similarity and joint angle-to-angle correlation) and both motor module similarity (**A**) and clinical outcomes (**B**). These results would suggest that improving the neuromuscular coordination under an isometric condition would improve kinematic coordination during a dynamic condition. In addition, following motor-module guided exercise, improvements in clinical score may be related to the transfer effect of neuromuscular coordination-guided exercise.*

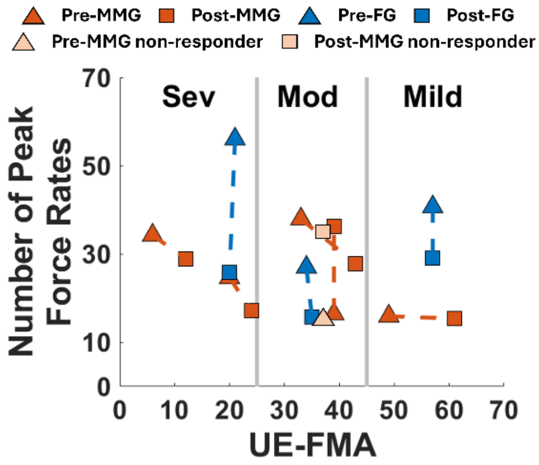


***Supplementary Figure 5.*** *Kinetic Analyses. Force profiles were sampled at 1000 Hz during the 54T isometric assessment. The force components (Fx, Fy, and Fz) were smoothed with cut-off frequency of 10 Hz using a 4th Butterworth low-pass filter. To evaluate the force control smoothness, the number of peaks in the force rate was calculated. We computed the first derivative of force trajectories from movement onset to offset for all directions and identified the number of peaks in the force rate. Then, these values were averaged across all targets for each participant. In the force-guided exercise group, the kinetics results showed a reduction in the number of peaks, showing that their force control was enhanced after training. Meanwhile, in the motor module-guided exercise group, improvement in the number of peaks was not observed.*
